# Top 50 cited articles on the treatment of chronic kidney disease: a bibliometric analysis

**DOI:** 10.64898/2025.12.15.25342322

**Authors:** Aryam S. Alshahrani, Norah A. Alodhaib, Mona G. Alhartani, Lamyaa M. Gharawi, Shoug A. Alshedokhi, Abdulaziz F. AlDafas, Zuhair A. Alabdullah, Mohamed A. Bassiony

## Abstract

**Introduction:** As the seventh greatest cause of premature mortality, chronic kidney disease (CKD) affects over 850 million people worldwide and poses a public health concern. Despite extensive research on CKD, it is still unclear which studies have had the greatest impact on treatment practices. Bibliometric analysis provides an evidence-based approach to identify key research trends, priorities, and global disparities.

This study aimed to determine the 50 most referenced publications on CKD treatment, examining citation patterns, thematic clusters, evidence quality, and global research distribution.

**Methods:** A comprehensive search of the Web of Science Core Collection was performed using the terms “Chronic Kidney Disease” AND “Treatment.” English-language publications involving human CKD treatment were included. After screening in Rayyan, data were extracted on study type, evidence level, treatment modality, and authorship. Analysis proceeded through four steps: (1) data cleaning and standardization; (2) descriptive bibliometrics of publication trends, core journals, and productivity indicators; (3) citation analysis using h-index–type metrics excluding self-citations; and (4) science mapping via VOSviewer and Bibliometrix to visualize keyword co-occurrence networks. Descriptive statistics summarized study characteristics.

**Results:** Randomized controlled trials accounted for only 16% of the top 50 papers (2000–2022), while narrative reviews comprised 42% and review articles 16%. Key subjects included SGLT2 inhibitors, metabolic therapies, and renin-angiotensin-aldosterone system blockade. Research output was concentrated in high-income regions, with limited contribution from low- and middle-income countries. Only 24% of publications achieved Level 1 evidence.

**Conclusion:** The literature on CKD treatment is geographically imbalanced, centralized, and dominated by narrative evidence. Region-specific bibliometric evaluations and more rigorous multicenter trials are required. For Saudi Arabia, aligning locally generated data with international knowledge is essential to develop equitable, context-driven, and evidence-based CKD management strategies.

## Introduction

According to the Kidney Disease: Improving Global Outcomes (KDIGO) 2021–2022 guidelines, CKD is defined as an abnormality of kidney structure or function that persists for more than three months and has outcomes for overall health^(1,2)^. Efforts have been made over the past 20 years to define and classify CKD in order to refine care on a global scale. In 2002, the Kidney Disease Outcomes Quality Initiative (KDOQI) was the first to propose a classification of CKD^(3)^. This work subsequently grew into the worldwide KDIGO framework (2012–2022), which added the cause-glomerular filtration rate–albuminuria (C–G–A) classification to the CKD stages and is now standard^(4)^. The disease is linked to premature mortality and frequent hospitalizations, exacting a heavy toll on the healthcare system^(5,6)^.

Chronic kidney disease (CKD) is an escalating global health concern, ranking as the 7th leading cause of early mortality. It is a worldwide burden that currently affects 850 million people, including low-and middle-income countries (LMICs), where access to consistent healthcare is often restricted.

Moreover, CKD is a major contributing factor to cardiovascular disease, further increasing the risk of morbidity and mortality^(5)^. The clinical treatment of chronic kidney disease (CKD) requires sustained management and cost-heavy care. Core treatment modalities, such as dialysis, medication-based treatment, and organ replacement therapy, are economically demanding and unevenly distributed globally. Despite long-term therapeutic intervention, progressive renal decline persists in a substantial proportion of CKD patients^(6)^.

Despite the abundant research conducted on CKD, identifying which studies are genuinely influential from the wide-ranging mass of publications remains considerably challenging. The overwhelming volume of literature contributes to uncertainty about the findings that truly influence clinical guidelines and patient management^(7)^. Regardless of the existence of several bibliometric studies in nephrology, they are mainly concentrated in dialysis or transplantation. Thus far, no study has yet addressed the broader landscape of CKD treatment research. A comprehensive assessment of original studies affecting CKD therapeutic strategies globally remains unexplored. Systematic analysis of research output and citation rates offers insights into which research findings impact the clinical community and rather notably policy makers, an aspect not addressed in a targeted manner^(8)^.

Citation analysis is a validated technique to detect influential research driving patient management and health policy. Although widely utilized in other specialties, it continues to be underused in CKD treatment, enabling the discovery of significant research and guiding evidence based decisions^(7)^.

Chronic Kidney Disease (CKD) research is growing globally; However, Saudi Arabia lacks citation analysis on CKD treatment. Local nephrology research output remains insufficient, and the relevance of internationally published studies to national medical use is limited. This gap restricts the development of localized, evidence-based decisions and healthcare policies^(9)^. Nearly 4.76% of Saudis have CKD, mainly at stage 3. Males and seniors are most affected. Diabetes, Hypertension, and obesity are primary risk factors. Obstacles include late-stage diagnosis, significant dialysis dependence, and restricted transplant choices^(10,11)^.

This study provides a systematic bibliometric assessment of CKD treatment literature, identifying leading research and revealing dominant trends in authorship, partnership, and global distribution. Through examining these factors, the study reveals the scientific landscape and identifies factors behind international therapeutic innovations. Essentially, the findings aim to integrate international research insights into the specific clinical and policy priorities in Saudi Arabia. This integration will support informed clinical decisions, improve distribution of healthcare resources, and influences future studies grounded in Saudi healthcare demands.

## Methods

### Search Strategy

The Web of Science core collection database was used to perform a thorough literature search with targeted Keywords (“Chronic Kidney Disease”) AND (“Treatment”). The search was not limited by the year of publication in order to guarantee a comprehensive collection of pertinent studies. Only English-language publications that particularly discuss the treatment of chronic kidney disease in humans were featured. A retrospective analysis was used as the study design, and it was carried out between July 3, 2025, and September 20, 2025. This study does not require ethical approval. The search approach was improved and optimized using a Boolean research design. Studies were initially arranged and assessed based on the quantity of citations they had received, giving preference to highly referenced publications that demonstrated a significant influence in the field. After screening, 474 of the 610 articles that were first found did not meet the inclusion criteria, whereas 136 of them did (Fig 1).

**Fig 1.**
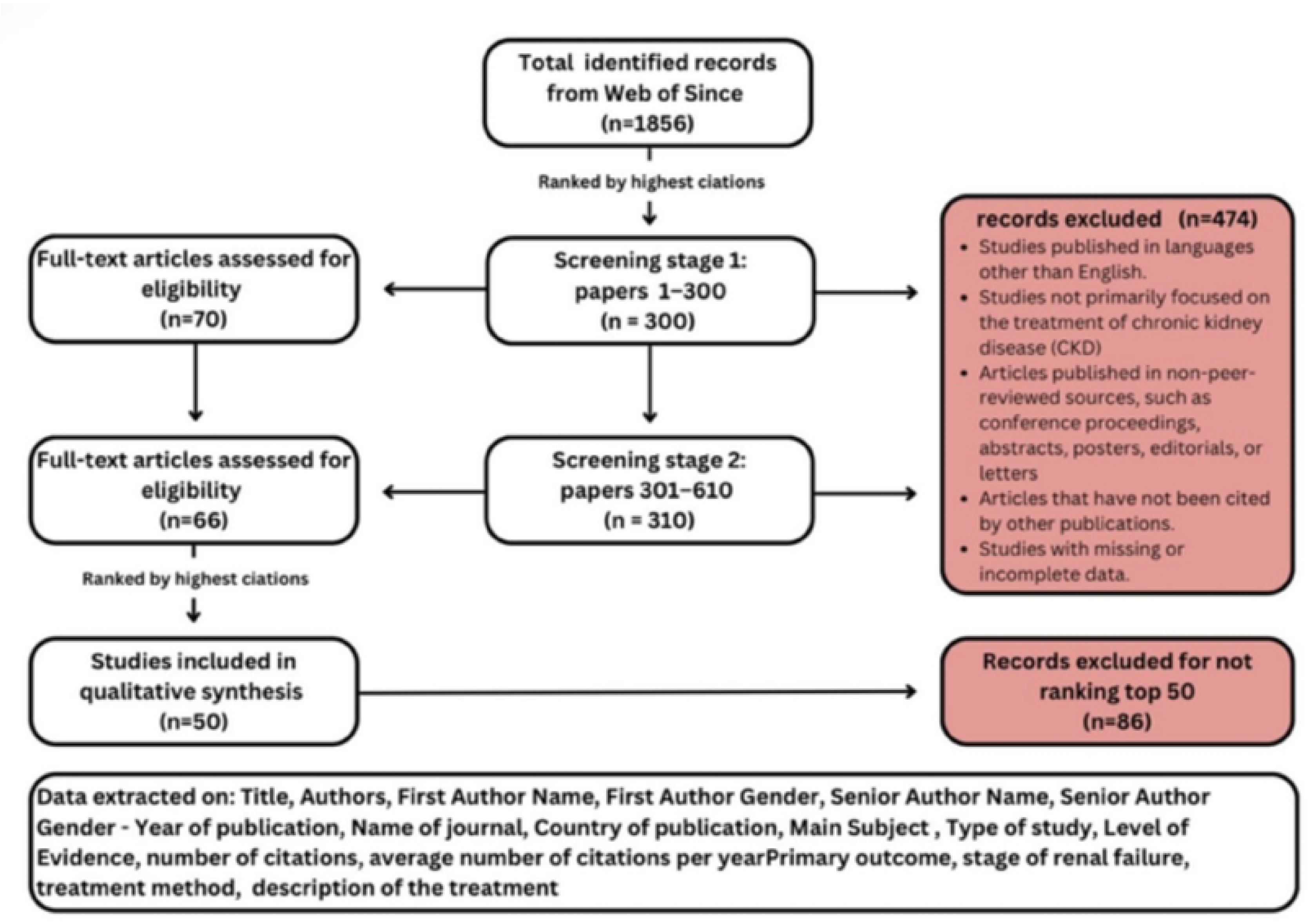
PRISMA-style flowchart outlining the study screening and selection process.

### Study Retrieval

Rayyan, a web-based tool for systematic review screening, was used to screen the literature. Three unbiased reviewers assessed the collected papers’ applicability by screening titles and abstracts according to predetermined inclusion and exclusion criteria. Studies were included if they were published in English without time constraints, had been referenced in other publications, featured human participants with chronic kidney disease (CKD), and specifically focused on CKD treatment. Excluded studies were those published in languages other than English, derived from non–peer-reviewed sources such as conference proceedings, abstracts, posters, editorials, or letters, those not referenced in other works, or studies with missing or incomplete data. After applying these criteria, three data collectors independently screened all retrieved papers and performed a full-text review. During the screening phase, any disagreements among the reviewers were resolved through discussion and consensus.

### Data Extraction and Management

A standardized form was designed to ensure consistency in collecting relevant data from each selected article. Three unbiased reviewers independently performed the data extraction. The retrieved data included several key variables, such as the title, authors, first author name, first author gender, senior author name, senior author gender, year of publication, journal name, and country of publication. Additional extracted information included the primary result, stage of renal failure, treatment approach (drug, surgery, transplantation, conservative, dialysis, complementary & alternative medicine, or others), type of study, level of evidence, primary subject, and treatment description. Any disagreements that arose during the data extraction process were resolved through discussion and consensus among the reviewers.

### Handling of Missing Data

To guarantee the accuracy and dependability of the findings, papers with missing or insufficient data were not included in the final analysis. This strategy was used to avoid potential bias resulting from incomplete datasets and to preserve uniformity across the included research.

### Statistical Analysis

Data analysis followed four linked steps. First, all records exported from the databases were merged and cleaned: duplicates were removed, incomplete entries were corrected, and author/institution names were standardized to ensure that productivity indicators reflected true output. Second, descriptive bibliometrics were generated—including annual publication trends, document types, core journals, and the most productive authors and countries—and were interpreted using classic bibliometric laws where relevant.

Third, impact was examined through citation analysis by calculating total and mean citations, identifying highly cited papers, and determining h-index–type indicators for key contributors; self-citations were checked to avoid inflated impact. Fourth, science-mapping techniques were applied to visualize the structure and evolution of the field. These included co-authorship analysis (to show collaboration patterns), co-citation and bibliographic coupling (to reveal intellectual bases and current research fronts), and keyword co-occurrence (to identify thematic clusters and emerging topics).

Visualizations were primarily produced using VOSviewer and Bibliometrix, with inclusion thresholds (e.g., minimum keyword occurrences) set to maintain interpretability. Where possible, analyses were stratified by time period to observe the evolution of themes and collaborations. Artificial intelligence tools were also used in the visualization process. Specifically, ChatGPT (GPT-5, OpenAI, 2025) was employed to generate figures based on the authors’ original data, and all outputs were reviewed and verified for accuracy and consistency with the results.

Complementary descriptive statistics were also employed to compile the characteristics of the top-cited studies. Continuous variables such as year of publication were summarized using the mean, median, and range. Categorical data—including primary subject, research type, and treatment strategy (pharmacological, surgical, transplantation, conservative, and other approaches)—were summarized using frequency distributions. The level of evidence was reviewed to identify temporal trends in research quality and influence. Although no formal bias assessment was conducted, a qualitative appraisal of treatment descriptions and primary outcomes was performed as part of the critical examination of each publication to provide insight into emerging research directions in the treatment of chronic kidney disease.

### Language editing and reference management

Artificial intelligence tools (ChatGPT, GPT-5, OpenAI, 2025; Scholar AI; and QuillBot) were used to assist with language editing and reference organization. All final content was reviewed and approved by the authors.

### Ethics statement

This study did not involve human participants, patient data, medical records, tissue samples, or any identifiable information. Therefore, institutional review board (IRB) approval and informed consent were not required.

## Results

### Top-Cited Literature on Chronic Kidney Disease (CKD) Treatments: Citation Patterns and Thematic Foci (2004–2022)

The corpus demonstrates a right-skewed citation distribution driven by a limited number of highly influential syntheses and clinical trials. The 2012 cluster, led by Wyld et al.’s utility-based quality-of-life meta-analysis (392 total citations), marked the field’s early shift toward incorporating health-economic outcomes alongside clinical efficacy. A second surge in citations occurred between 2019 and 2022, corresponding to studies on cardio-renal–metabolic therapeutics and implementation research, notably involving SGLT2 inhibitors (e.g., Mende 2022)^(22)^, non-steroidal MRAs/finerenone (Agarwal et al. 2022; Rossing et al. 2022)^(31)^, and bariatric surgery (Cohen et al. 2020; Docherty & le Roux 2020)^(15)^. These recent publications exhibit high annual citation rates, reflecting the rapid diffusion of practice-changing evidence, whereas older mechanistic or narrative works (e.g., oxidative stress, fibrosis) f a more linear citation pattern over time.

Methodologically, the top-cited studies integrate evidence synthesis (systematic reviews/meta-analyses, and guideline-linked reviews), randomized and subgroup analyses (FIDELIO/FIGARO-DKD-adjacent), and health services/economic evaluations (e.g., early referral and DAPA-CKD cost-effectiveness analyses). This diversity reflects translational continuity from mechanisms (fibrosis, endothelin, inflammation) to therapeutic advances (RAAS blockade, MRAs, SGLT2is, patiromer) and value-based care dimensions (referral timing, targets, economics).

Notably, complementary approaches—traditional Chinese medicine (TCM), herbal interventions, and microbiome modulation via resistant starch—form a distinct secondary stream, indicating geographic and paradigmatic diversity, although their citation velocity remains lower than that of mainstream pharmacotherapy and trial-aligned reviews.

This therapeutic diversity is illustrated in Fig 2, which displays cluster patterns of treatment categories across the corpus. Pharmacologic and conservative therapies form the largest and most interconnected clusters, underscoring their predominance and frequent co-occurrence in CKD literature. Procedural and renal replacement interventions appear as smaller but distinct clusters, while alternative and complementary modalities are positioned peripherally, reflecting a specialized yet expanding research stream within the global CKD treatment landscape.

**Fig 2.**
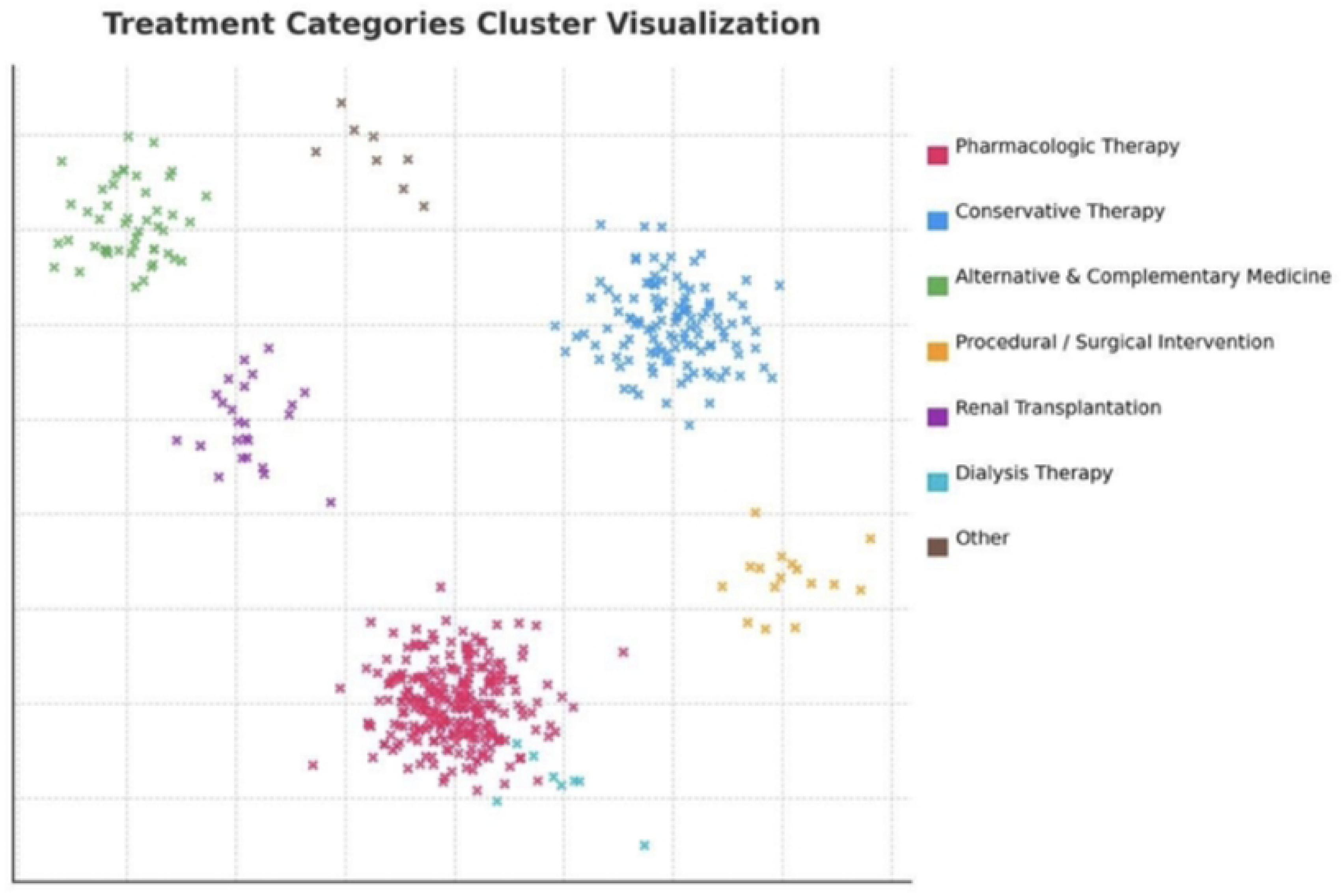
Cluster visualization of treatment categories across the top-cited corpus.

Table 1 lists the top 50 cited articles, detailing their authors, total citation counts, and average number of citations per year.

**Table 1:**
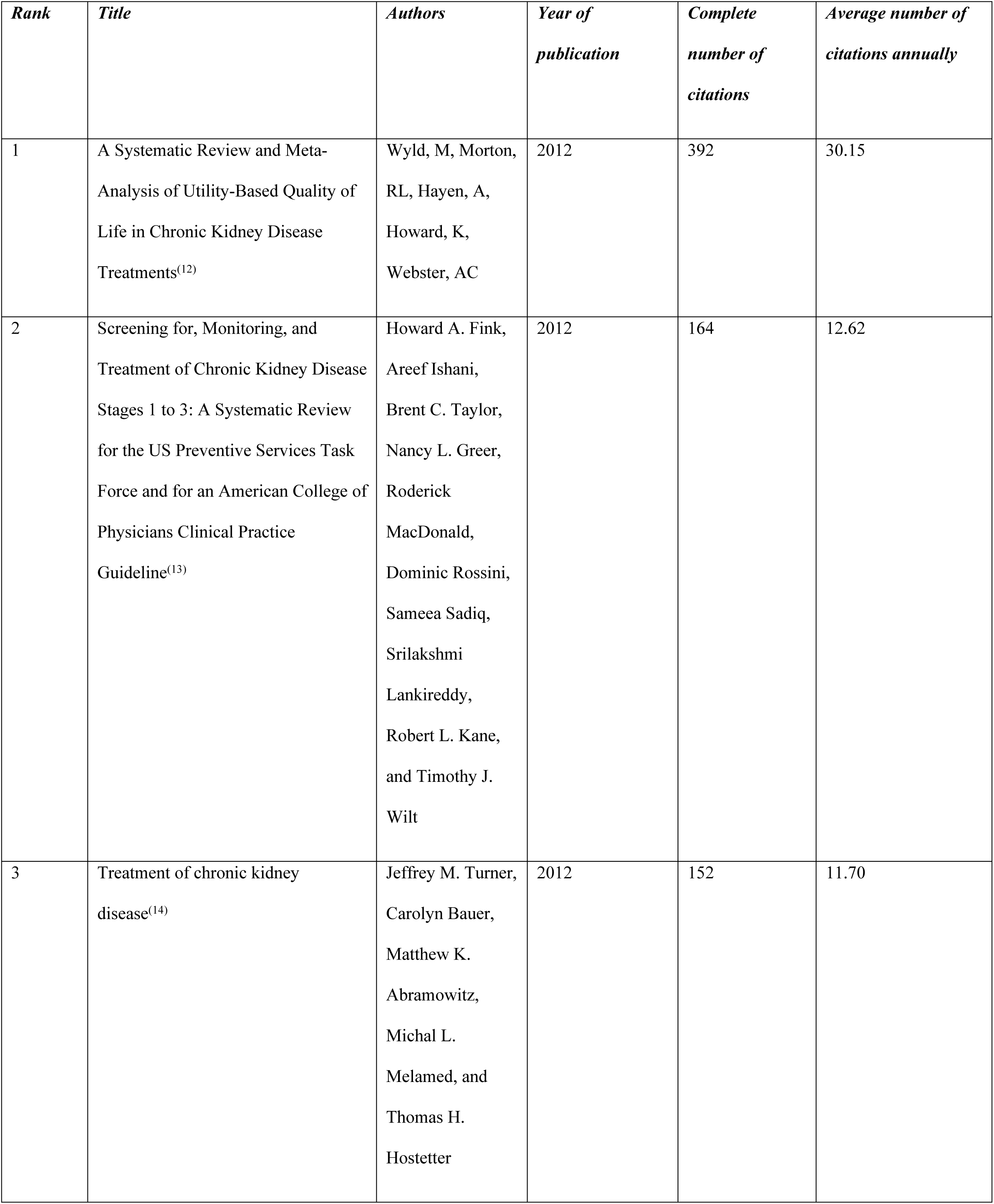
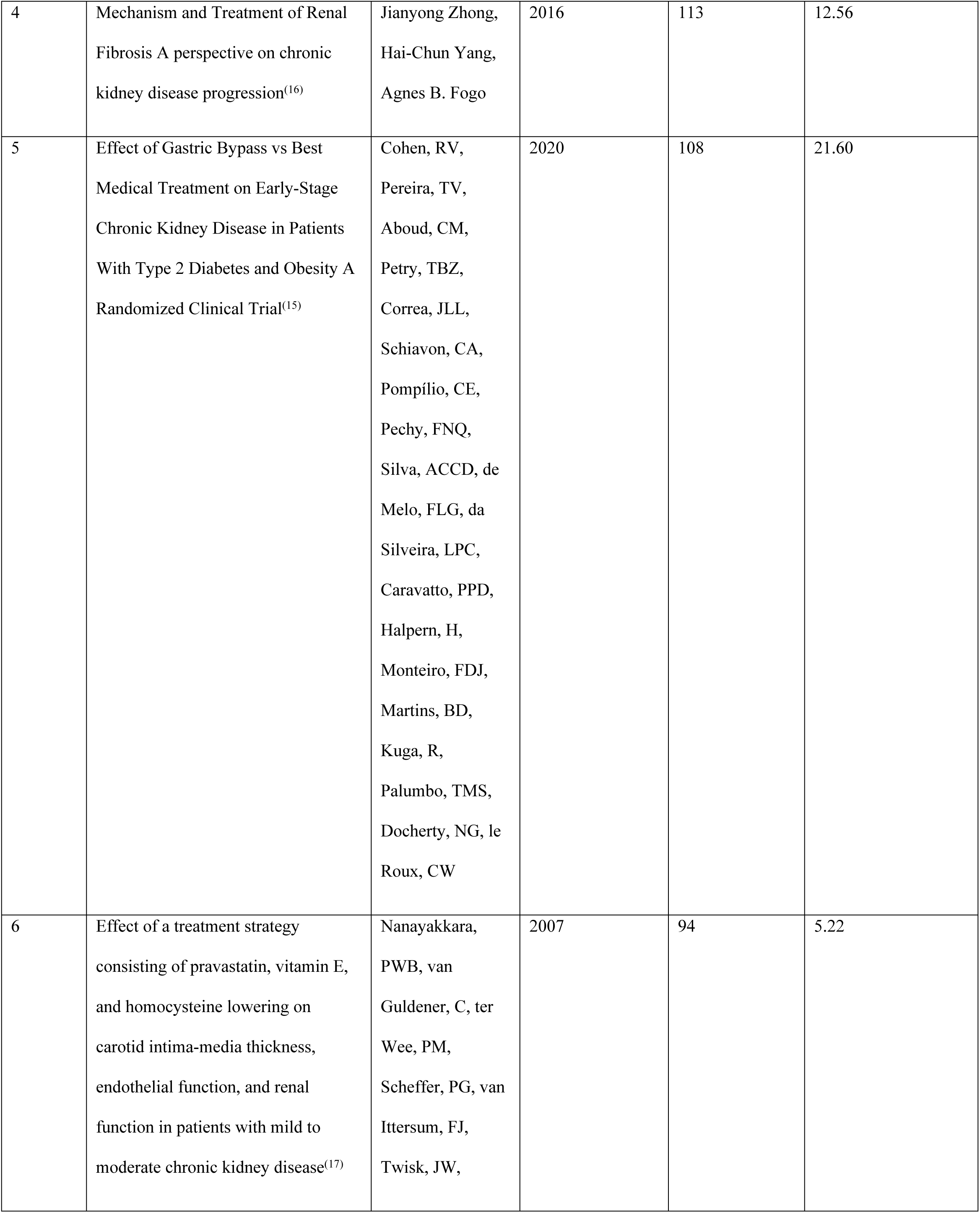
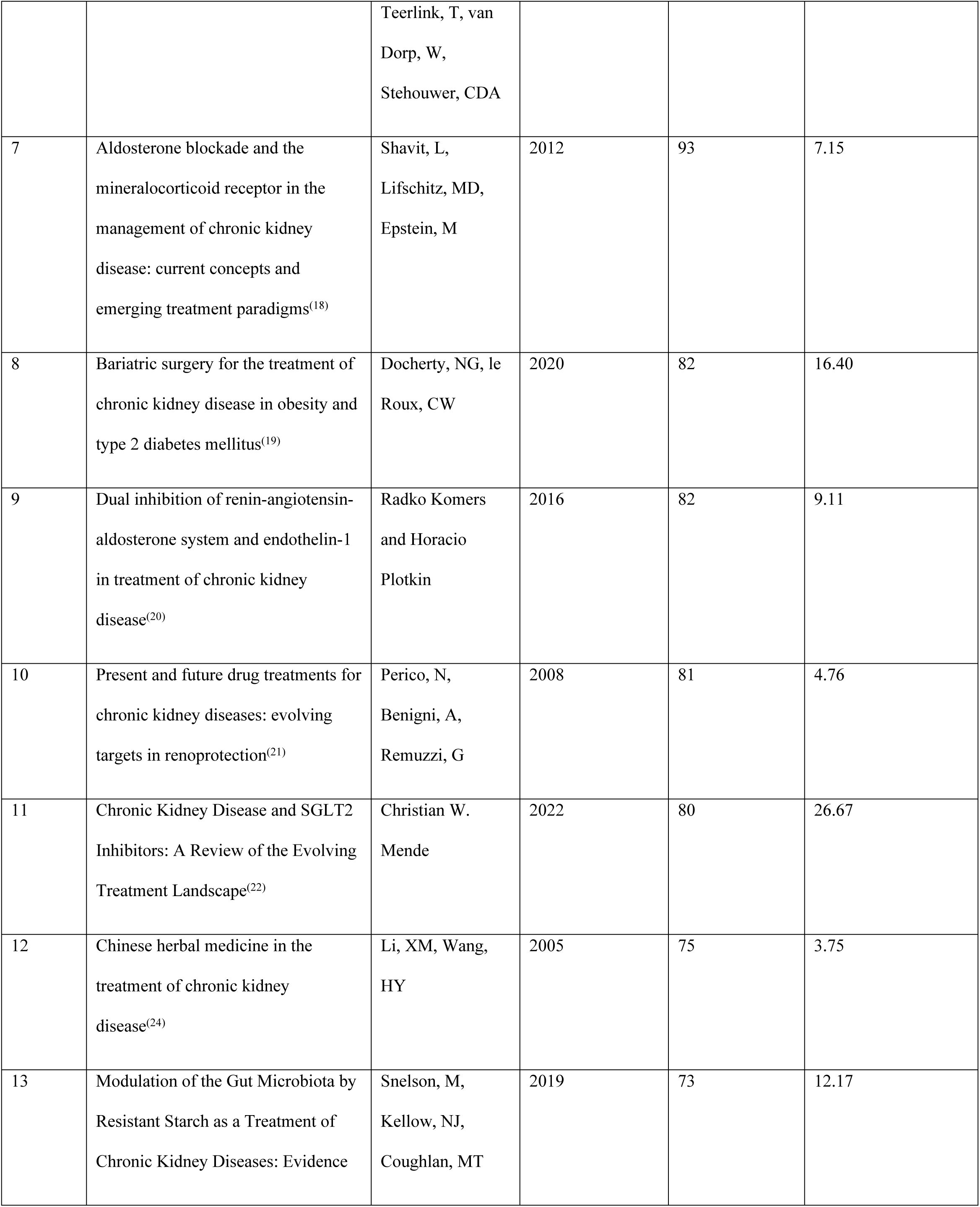
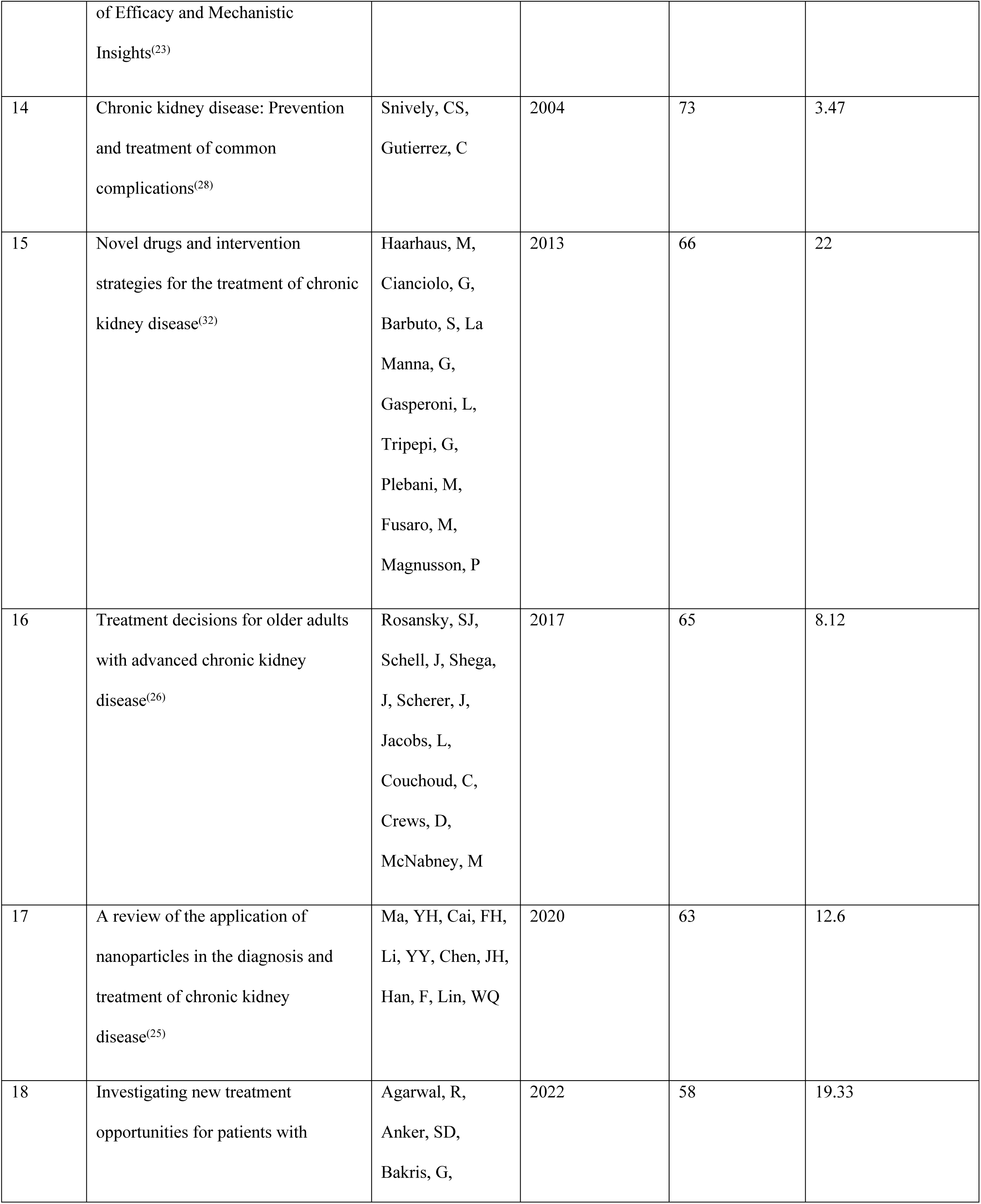
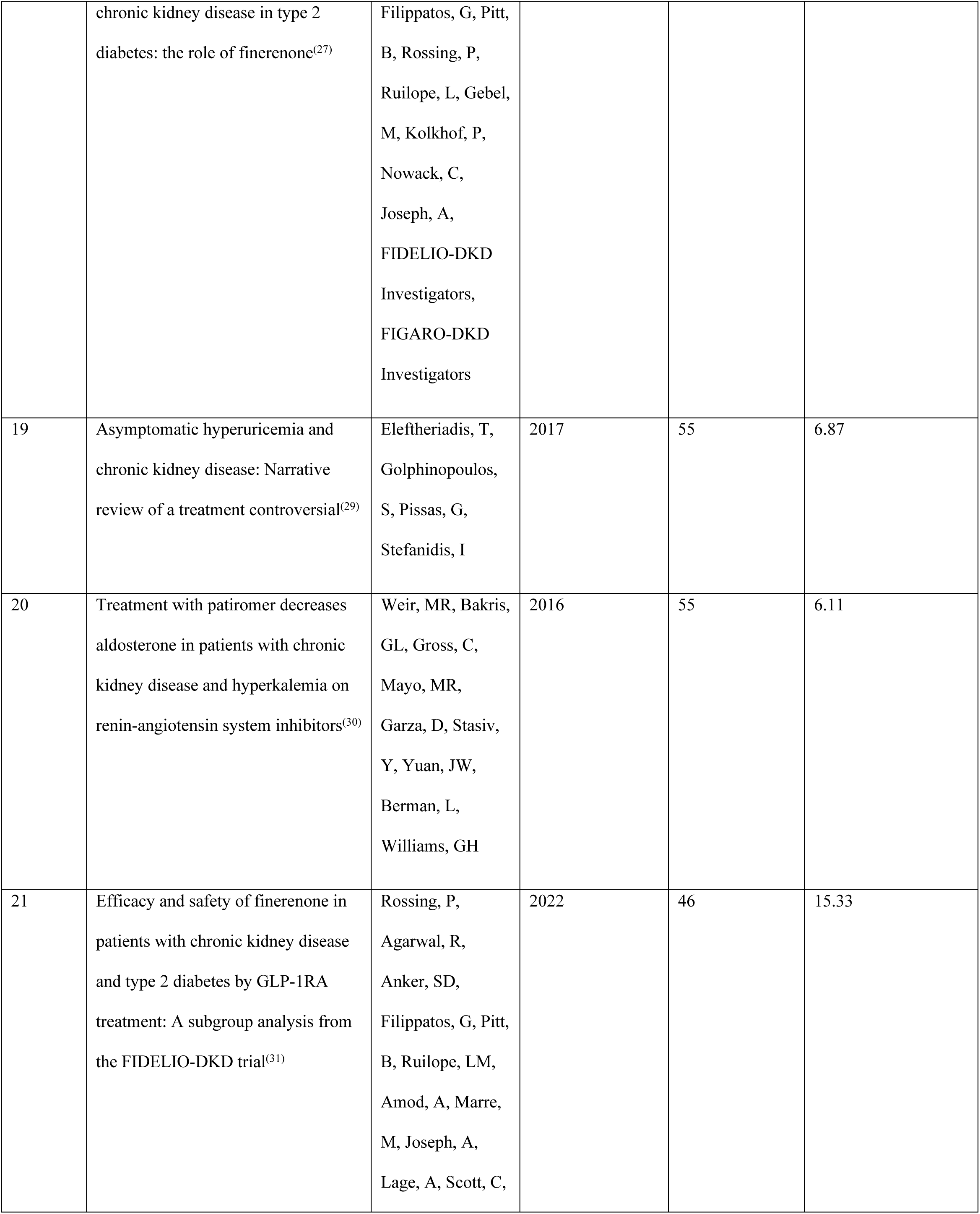
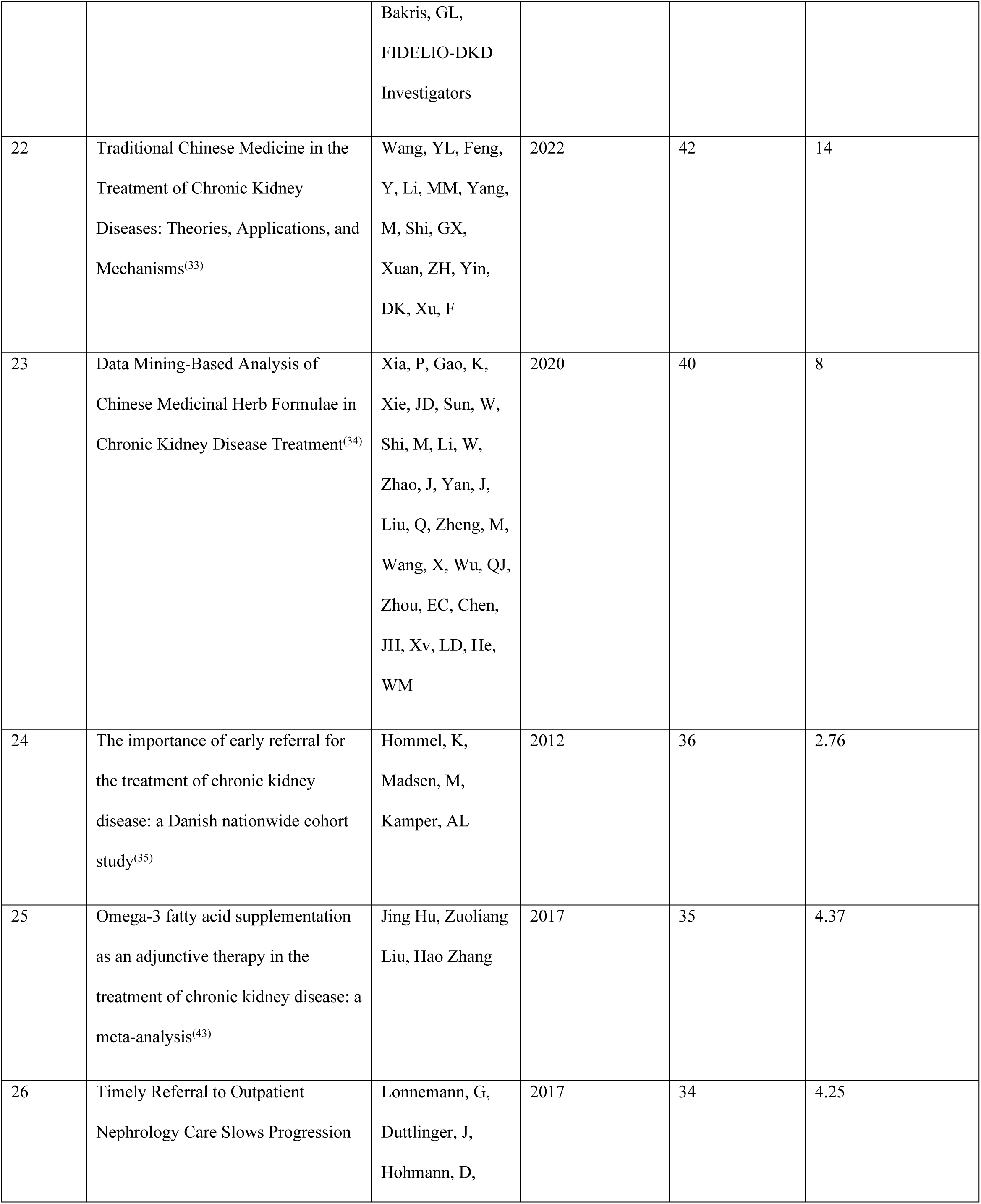
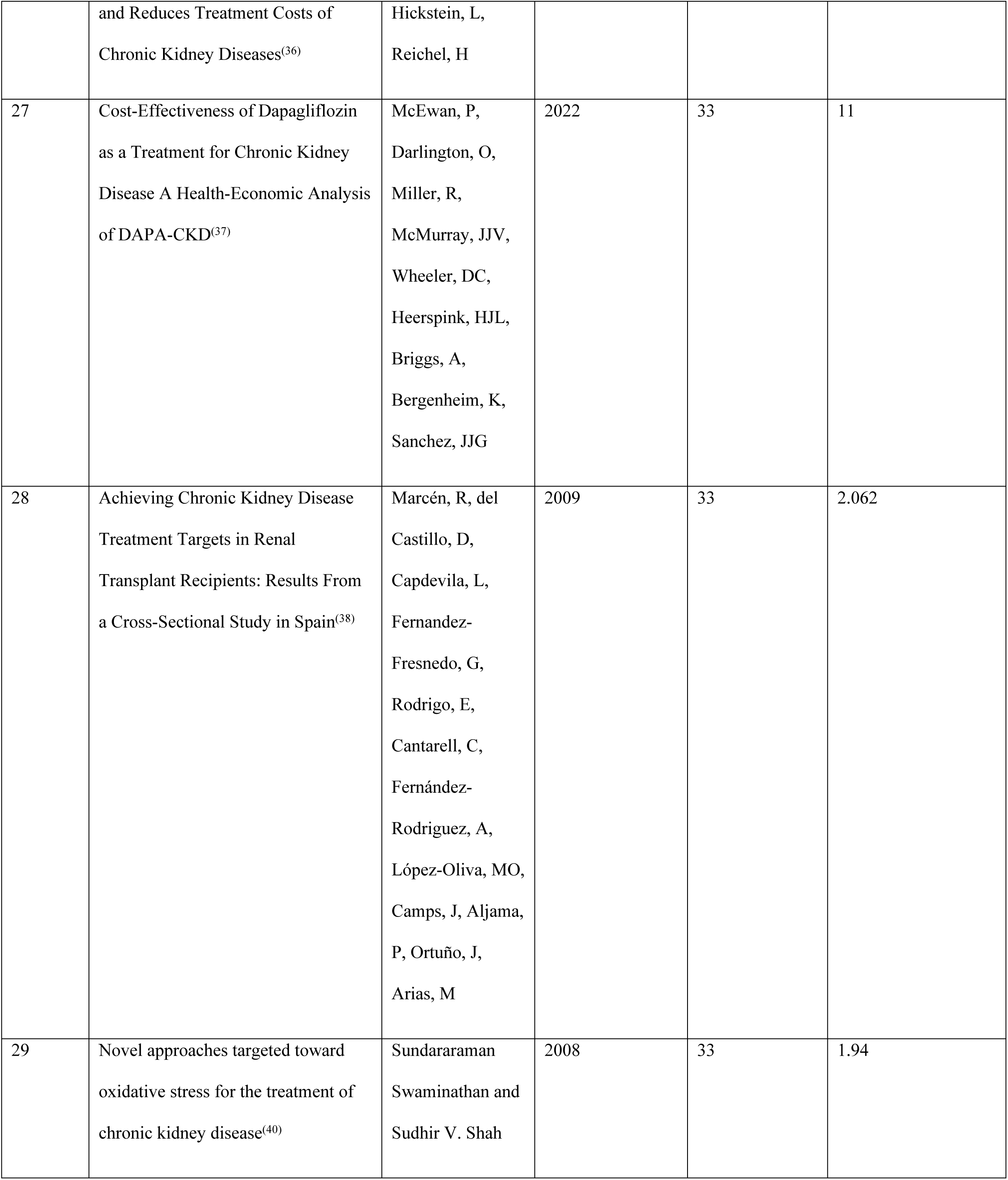
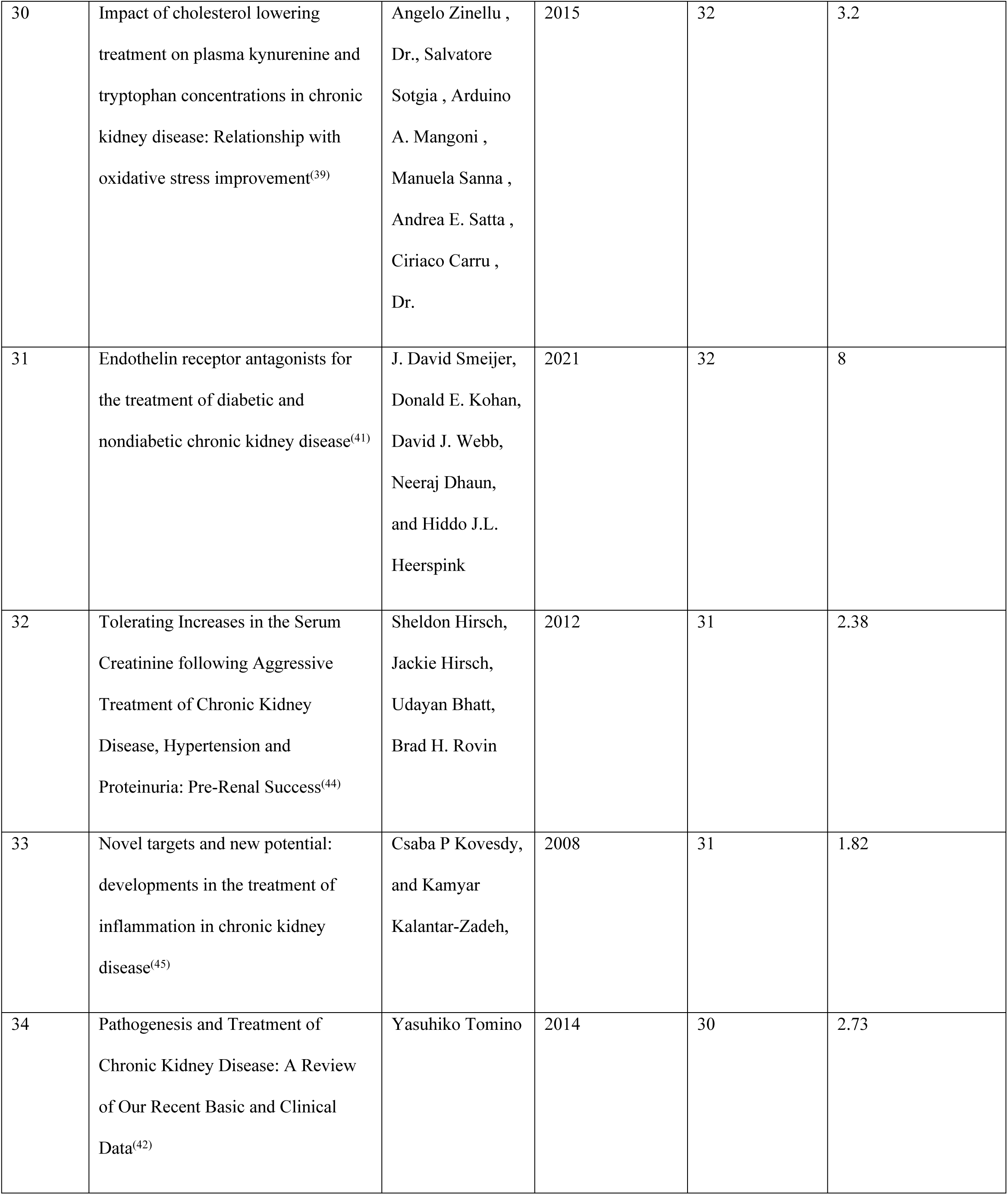
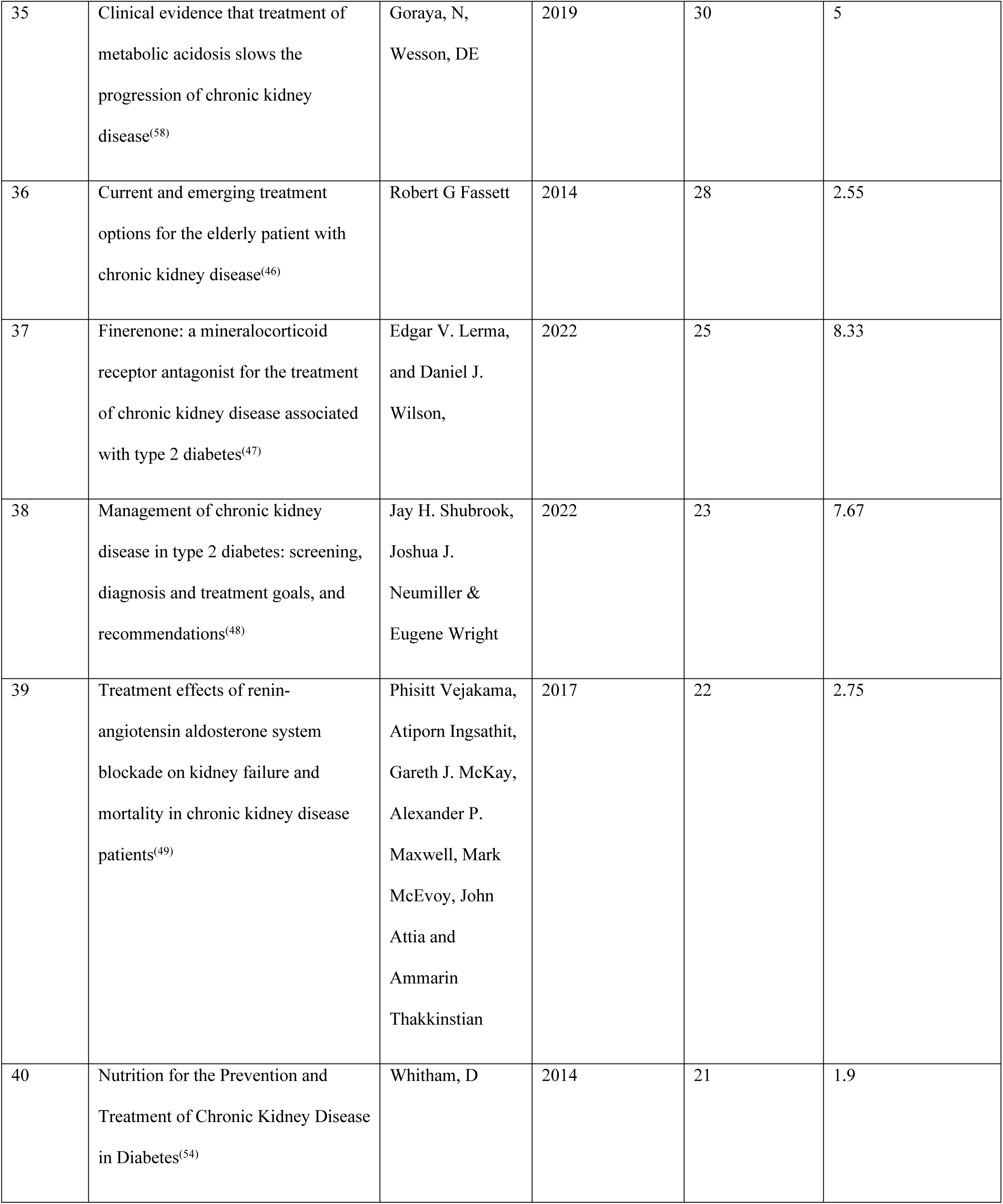
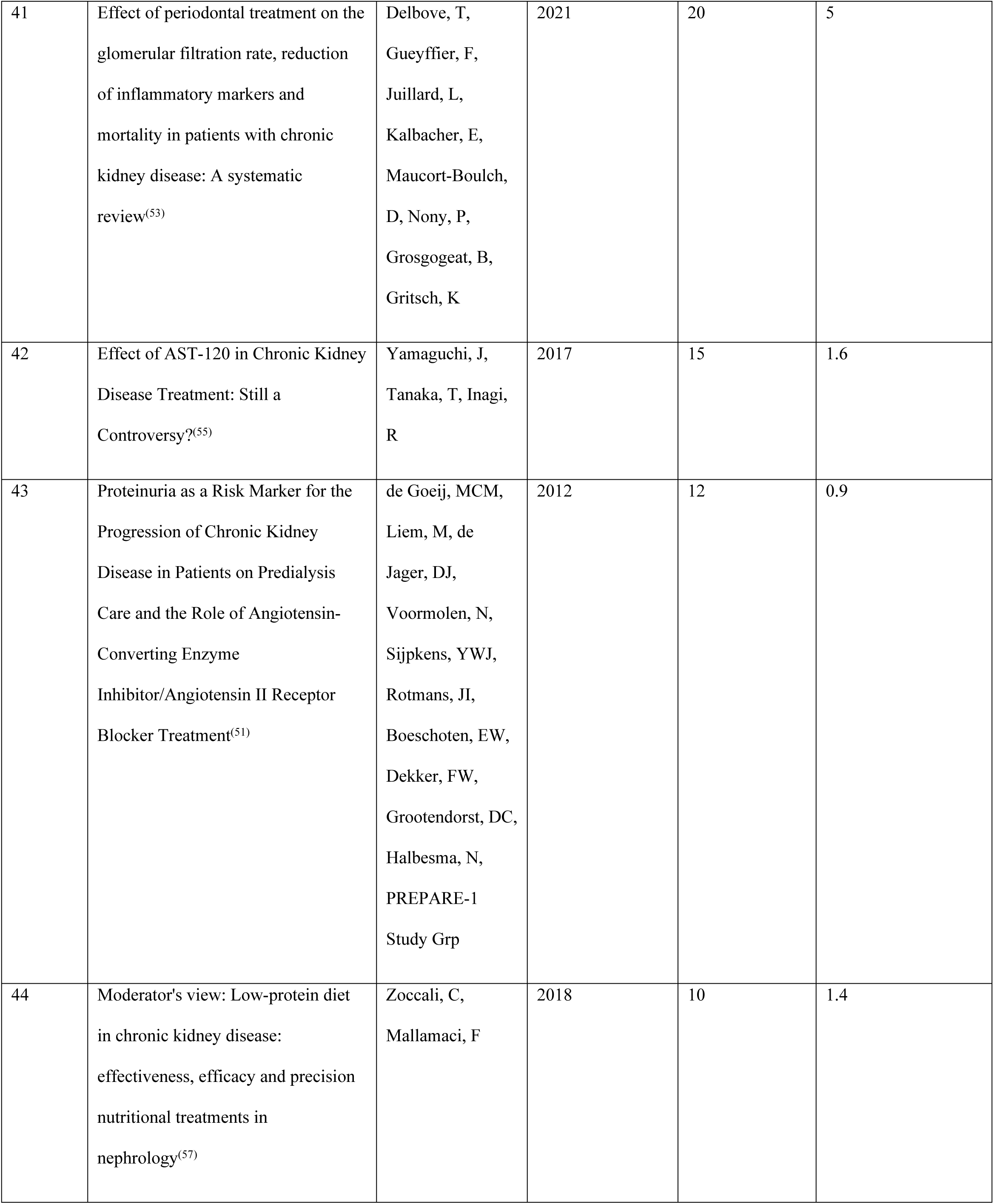
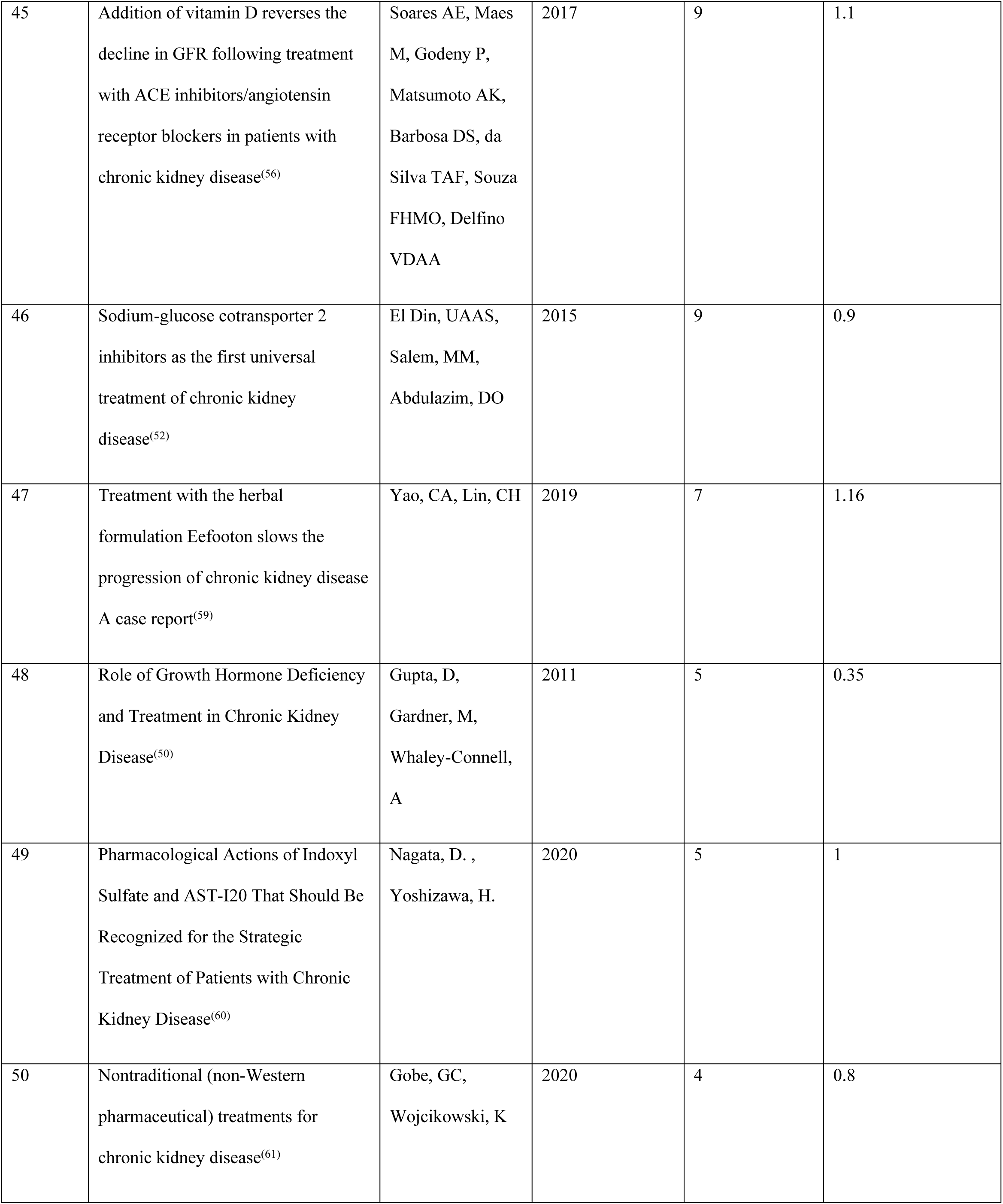
Top 50 Cited Literature on Chronic Kidney Disease (CKD) Treatments.

### The keyword co-occurrence

The keyword co-occurrence analysis of chronic kidney disease (CKD) research reveals a densely interconnected and multidisciplinary scholarly landscape (Fig 3). At the center of the network, terms like “chronic kidney disease,” “glomerular filtration rate,” “mortality,” “prevalence,” “epidemiology,” and “population” dominate, reflecting the central focus on disease burden quantification and progression metrics in CKD literature. Surrounding clusters highlight distinct yet interrelated research themes. The green cluster emphasizes public health and epidemiological dimensions, encompassing *awareness*, *intervention*, and *classification*. The blue and purple clusters focus on therapeutic and clinical management aspects including *dialysis*, *therapy*, and comorbidities like *cardiovascular disease*, *diabetes*, and *stroke*. In contrast, the red and yellow clusters explore mechanistic and pathophysiological underpinnings, featuring *oxidative stress*, *inflammation*, *biomarkers*, and *genetic* factors. Emerging topics such as *gut microbiota*, *nutrition*, and *adipose tissue* suggest growing interest in metabolic and systemic contributors to CKD progression. The network also reveals methodological trends, with frequent references to *trials*, *guidelines*, and *evidence-based recommendations*, signaling the field’s advancement toward standardization and global best practices. Overall, the map illustrates that CKD research spans molecular biology to public health, with core emphasis on understanding disease prevalence, risk stratification, and outcomes across diverse global populations.

**Fig 3.**
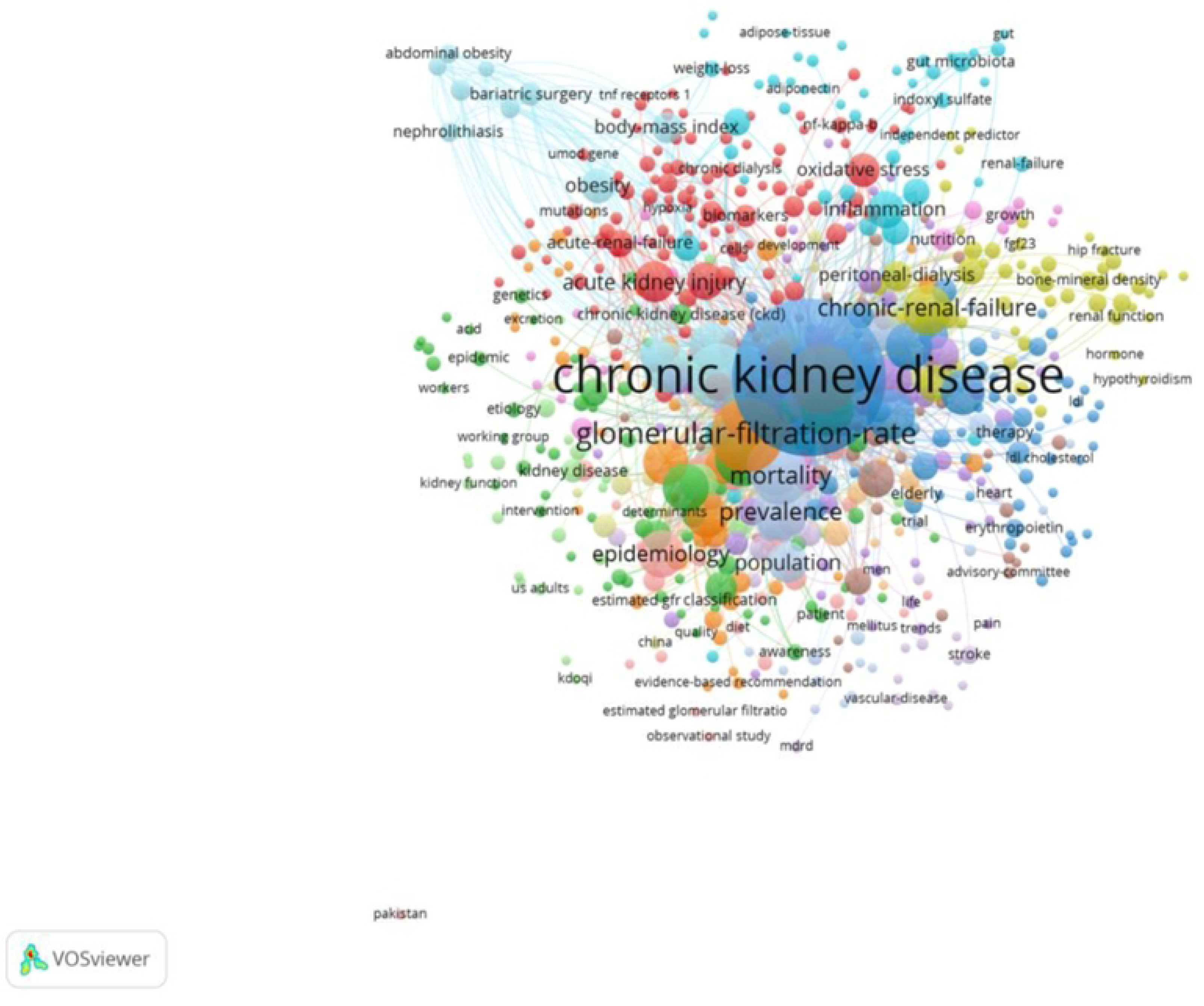
Keyword co-occurrence network illustrating dominant CKD research themes.

### The citation co-occurrence

Analysis of author collaborations within the top-cited CKD treatment literature reveals a relatively sparse network composed primarily of dyadic first–senior author ties. Within this structure, several recurrently recognized investigators emerge as local hubs through their connections to influential first authors.

A notable collaboration includes Webster, A.C., paired with Wyld, M., representing evidence-synthesis and management-focused contributions to CKD care. A second methodological axis is formed by Wilt, T.J. with Fink, H.A., reflecting trial- and guideline-oriented research on screening and treatment strategies. Hostetter, T.H. with Turner, J.M. anchored authoritative reviews bridging mechanistic and clinical perspectives on CKD pathophysiology, while Fogo, A.B. with Yang, J.Z. Contributed to fibrosis and progression biology. Metabolic and procedural approaches are represented by Roux, C.W.L. alongside Cohen, R.V., highlighting bariatric and surgical interventions within advanced CKD populations. Additional recurrent pairings—Remuzzi, G. with Perico, N.; Epstein, M. with Shavit, L.; Plotkin, H. with Komers, R.; Stehouwer, C.D.A. with Nanayakkara, P.W.B.—highlights the field’s diversity of mechanistic and clinical strands across RAAS modulation, hemodynamic pathways, and cardiometabolic risk.

Overall, the collaboration landscape portrays a field anchored by influential senior investigators working with distinct first authors across topics spanning evidence synthesis, clinical trial methodology, mechanistic underpinnings of fibrosis and progression, and emerging metabolic/surgical strategies—signaling a mature yet distributed authorship landscape in nephrology.

### Decade distribution

Among the 50 most cited articles on the treatment of chronic kidney disease (CKD), the 2010–2019 period represents the most productive decade, contributing more than half of the publications (n = 27; 54.0%) (Table 2; Fig 4). This period reflects a substantial rise in research activity and global attention toward evidence-based treatment of CKD. The 2020–2022 period (n = 16; 32.0%) has also shown considerable productivity despite being an incomplete decade, indicating ongoing and growing interest in the field. This recent surge likely aligns with the renewed focus following the development of the updated KDIGO clinical practice guidelines for CKD treatment in 2021–2022. In contrast, the 2000–2009 period (n = 7; 14.0%) contributed fewer high-impact publications, possibly representing early groundwork that set the stage for later advances. Overall, this pattern highlights a clear expansion of scientific output over time, peaking in 2010–2019 and maintaining strong momentum into 2020–2022, likely driven by the increasing global burden of CKD and evolving clinical standards.

**Fig 4.**
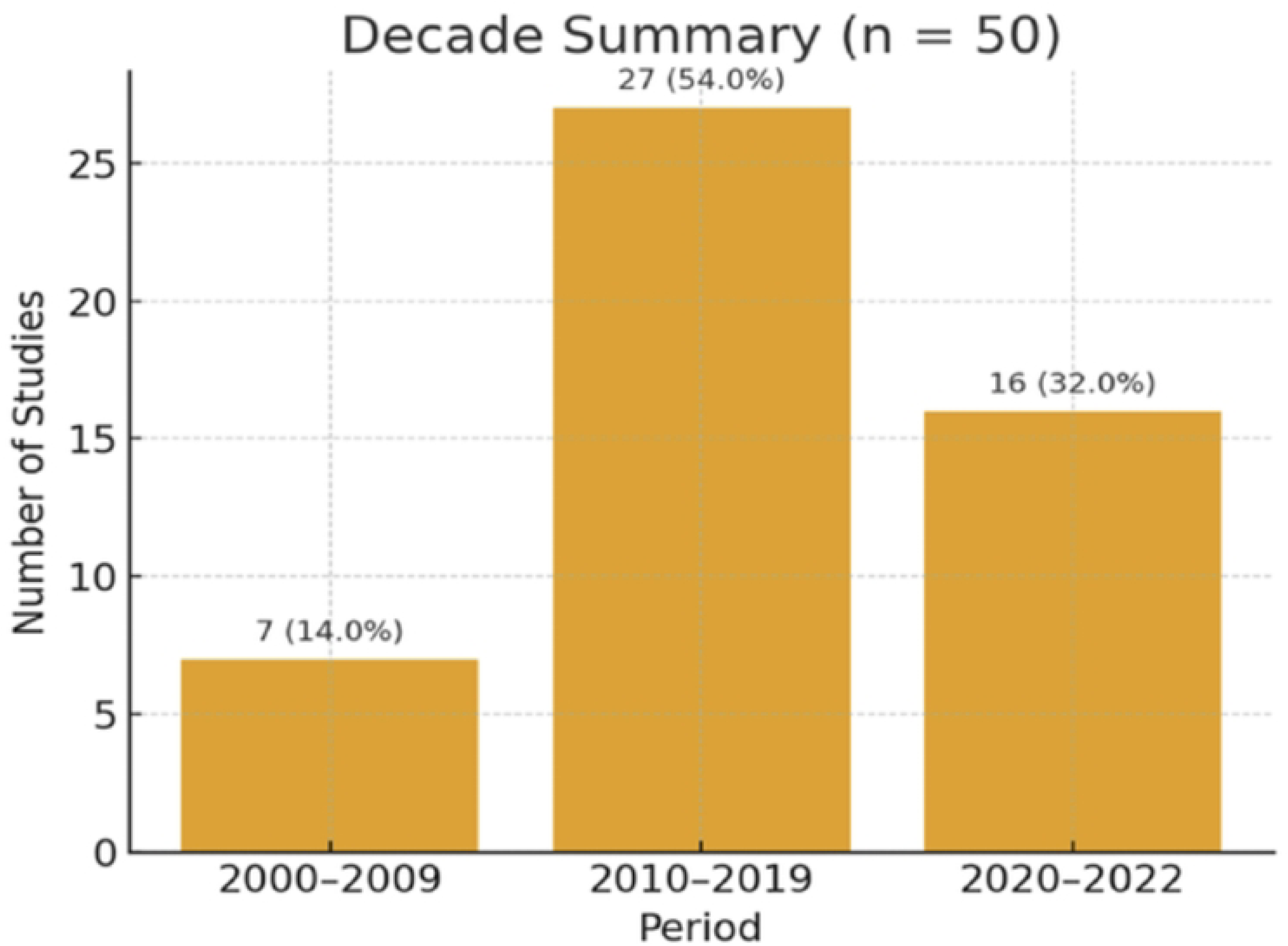
Decadal distribution of highly cited CKD treatment studies.

**Table 2:**
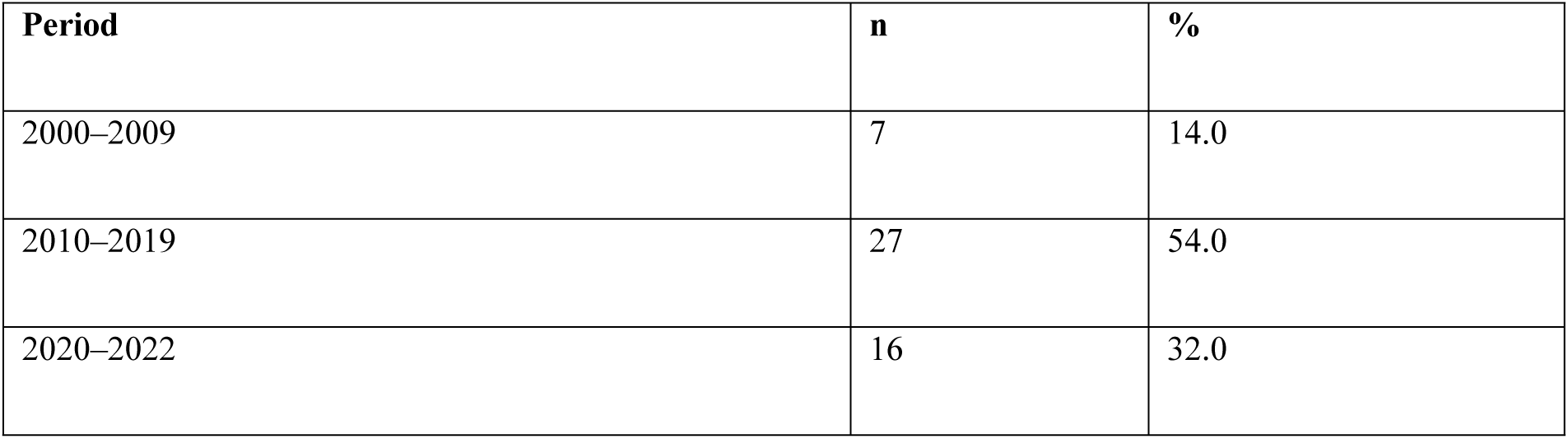
Decade Distribution of the 50 Most Cited CKD Treatment Articles.

### Publishing journals distributions

The distribution of journals publishing the most cited articles on the treatment of chronic kidney disease demonstrates a diverse and interdisciplinary research landscape, with 43 journals contributing to the field (Table 3). Most journals published only a single highly cited article; however, a few stood out as recurrent sources of influential research.

**Table 3:**
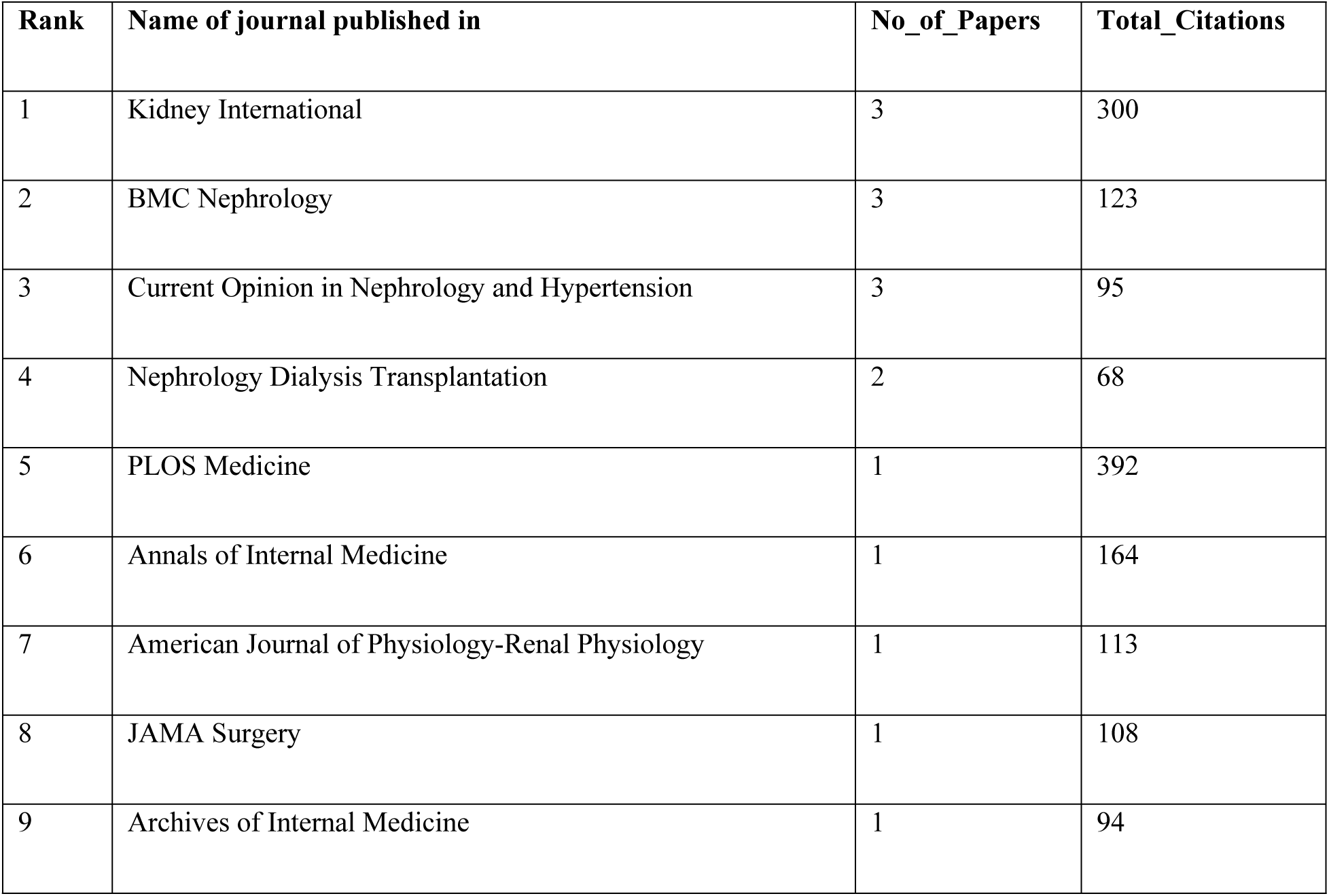
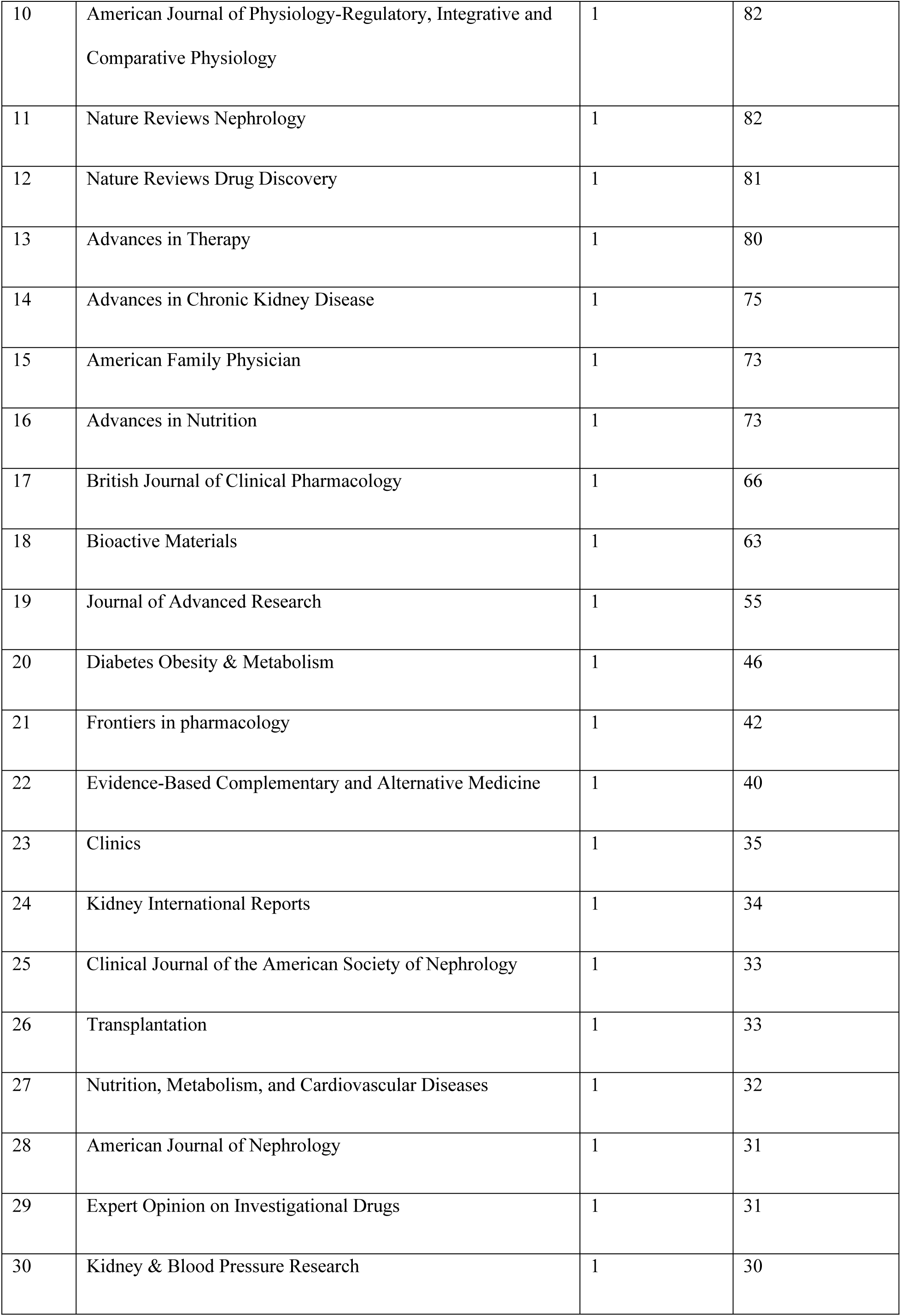
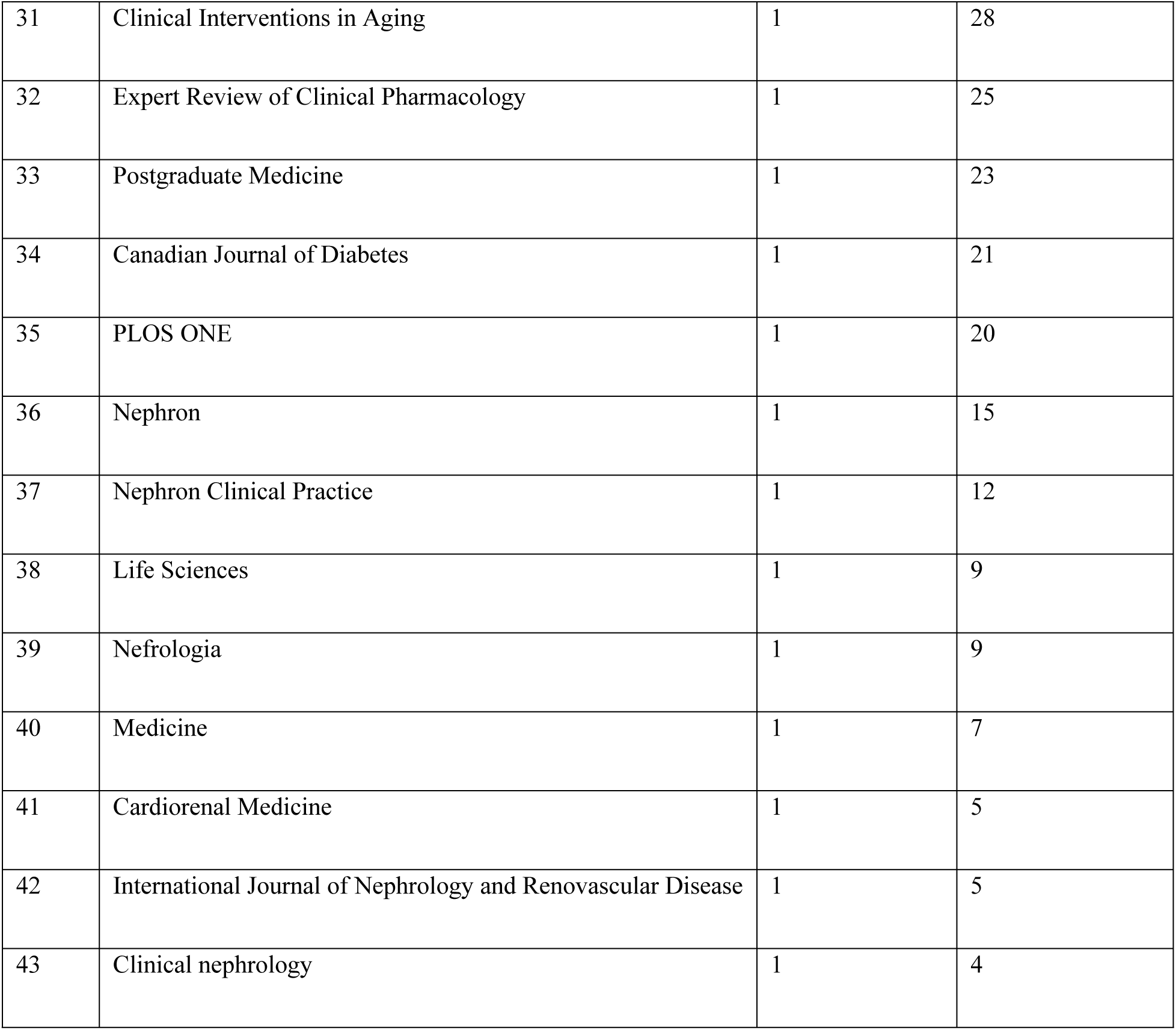
Journals Publishing the Top-Cited CKD Treatment Articles.

*Kidney International* ranked first, contributing three papers that collectively received 300 citations, underscoring its pivotal role as a leading source of high-impact nephrology research. Other recurrent contributors included *BMC Nephrology* and *Current Opinion in Nephrology and Hypertension*, each publishing three articles with total citations of 123 and 95, respectively. *Nephrology Dialysis Transplantation* followed with two papers and 68 citations, highlighting its continued relevance in disseminating advances in dialysis and transplant-related therapy.

Other high-impact general medical journals such as *PLOS Medicine* (392 citations) and *Annals of Internal Medicine* (164 citations) also featured prominently, despite each contributing only one article. Their presence highlights how publication in broad-scope, top-tier medical journals can lead to substantial visibility and academic influence.

Beyond traditional nephrology outlets, a diverse range of journals contributed to the citation landscape— including those focused on physiology, pharmacology, nutrition, surgery, and alternative medicine. Examples include the *American Journal of Physiology–Renal Physiology* (113 citations), *JAMA Surgery* (108 citations), and *Evidence-Based Complementary and Alternative Medicine* (40 citations). Notably, journals such as *Bioactive Materials*, *Advances in Nutrition*, and *Expert Opinion on Investigational Drugs* reflect the growing integration of biomaterials, nutritional science, and pharmacotherapy in nephrology-related research.

This broad distribution across specialized and interdisciplinary journals underscores the multifaceted nature of chronic kidney disease research. It reflects the integration of diverse scientific domains that collectively contribute toward improving renal function, patient outcomes, and overall quality of care.

### Publishing countries distribution

Regarding geographical context, the top-cited articles originated from 17 countries (Table 4; Fig 5). Research output was highly concentrated, with the United States leading (15/50; 30.0%), followed by China (5/50; 10.0%) and the Netherlands (5/50; 10.0%), which had the same number of publications. The United Kingdom (4/50; 8.0%) and Australia (4/50; 8.0%) also contributed an equal number of articles. Italy (3/50; 6.0%) and Japan (3/50; 6.0%) each produced three publications, while Egypt contributed two (2/50; 4.0%). Nine additional countries—Brazil, Canada, Denmark, France, Germany, Ireland, Spain, Taiwan, and Thailand—each produced one article (1/50; 2.0%).

**Fig 5.**
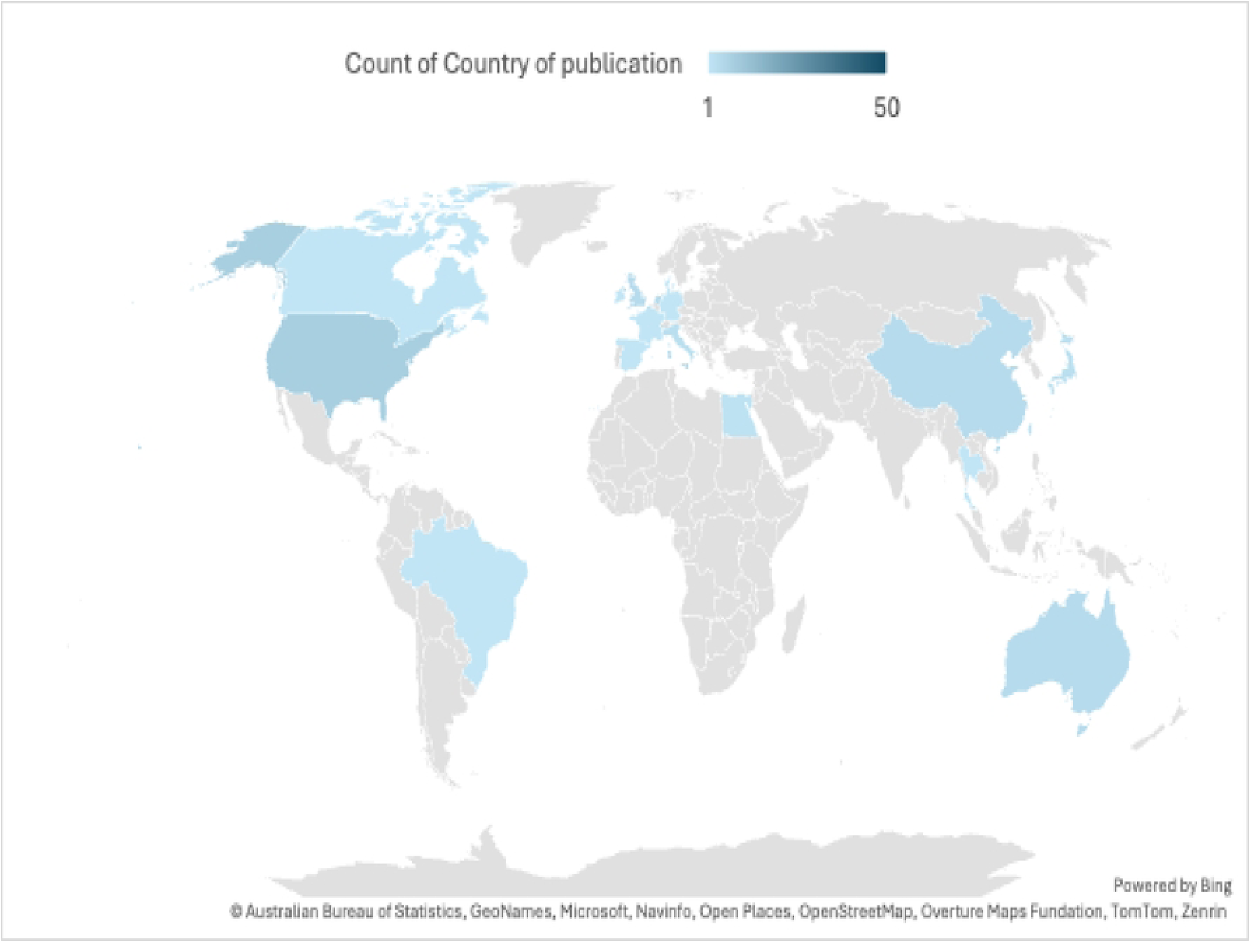
Geographic distribution of study settings among the top 50 most-cited CKD treatment papers.

**Table 4:**
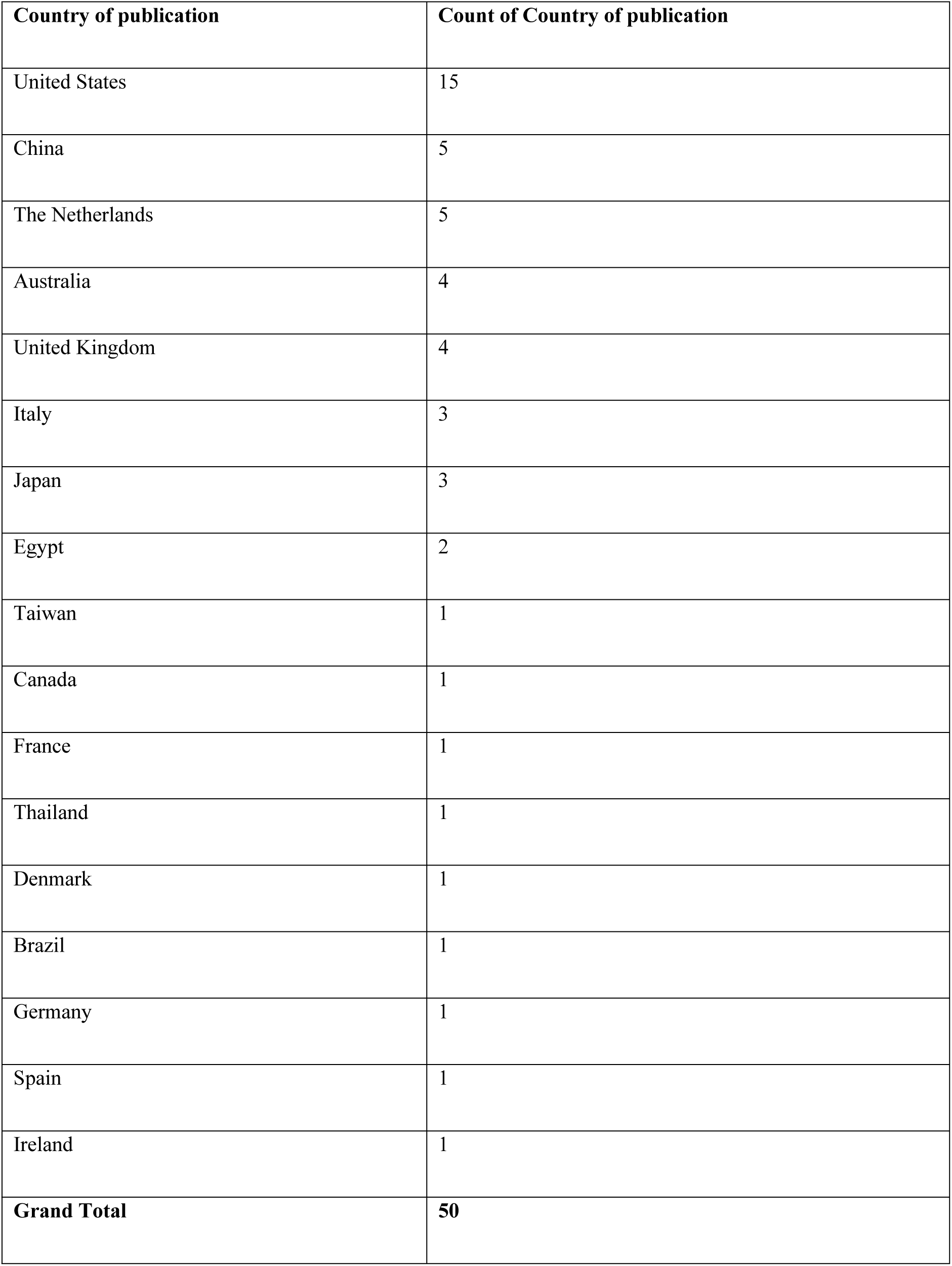
Distribution of Articles by Country.

When grouped by region, Europe accounted for 17 articles (34.0%), North America for 16 (32.0%), the Asia–Pacific region for 14 (28.0%), the Middle East/North Africa for 2 (4.0%), and Latin America for 1 (2.0%). Overall, the geographical distribution demonstrates broad international engagement in CKD treatment research but remains skewed toward high-income regions, with the top five contributing countries responsible for 33 of 50 articles (66.0%).

### Study design and level of evidence

Among the top 50 cited articles, review-type studies predominated (Table 5; Fig 6) with narrative reviews (21/50; 42.0%) and review articles (8/50; 16.0%) together comprising 58.0% of the total. Randomized controlled trials (RCTs) accounted for 8/50 (16.0%). Observational designs included retrospective cohorts (5/50; 10.0%), prospective cohorts (2/50; 4.0%), and cross-sectional studies (1/50; 2.0%). One case report was identified (1/50; 2.0%). Evidence syntheses, reported as separate pre-specified categories, included systematic reviews (2/50; 4.0%) and systematic reviews with meta-analyses (2/50; 4.0%).

**Fig 6.**
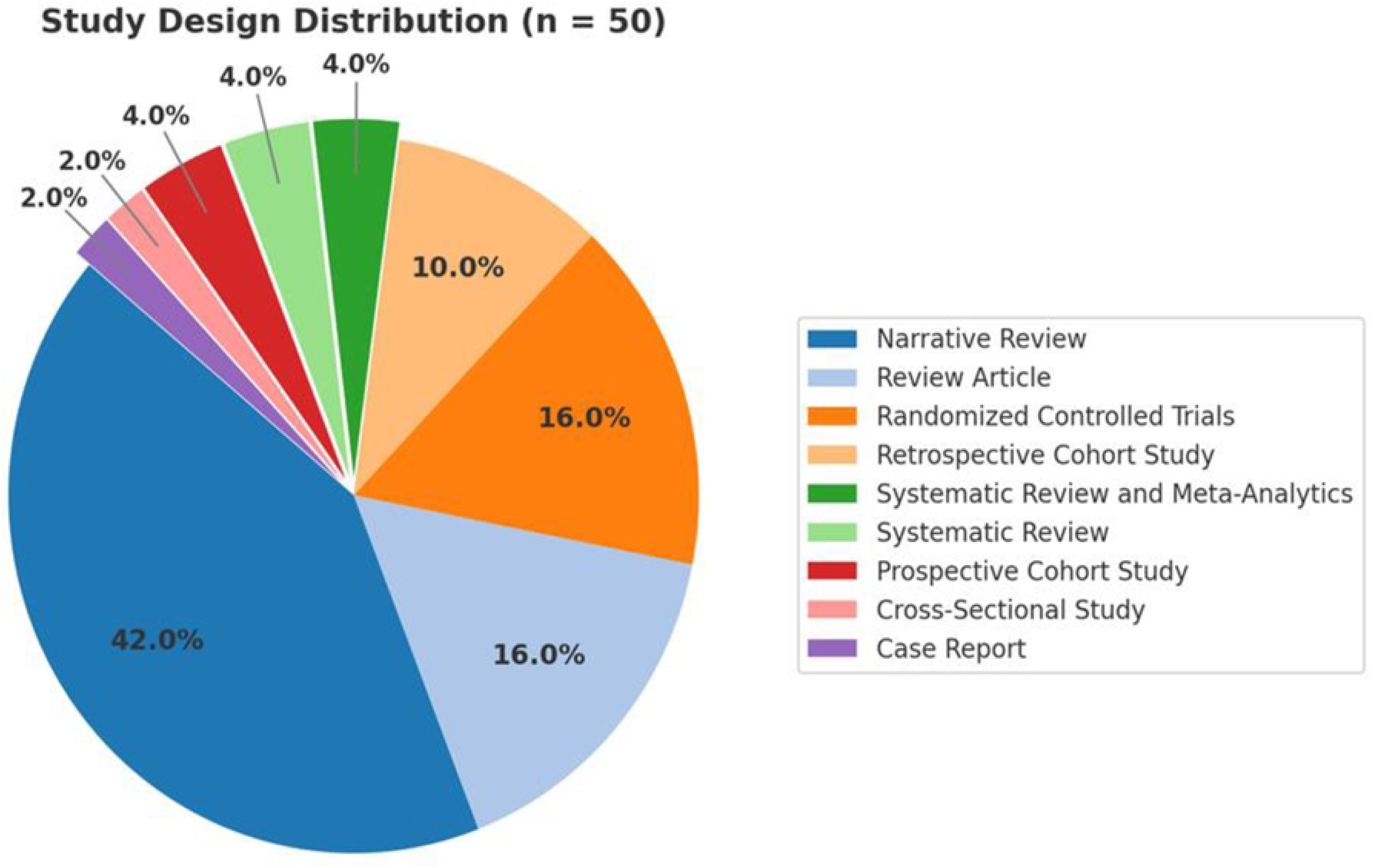
Study designs of the most influential CKD treatment publications.

**Table 5.** Study Design Distribution.

| **Study design** | **n** | **%** |
| --- | --- | --- |
| Narrative Review | 21 | 42.0 |
| Review Article | 8 | 16.0 |
| Randomized Controlled Trials | 8 | 16.0 |
| Retrospective Cohort Study | 5 | 10.0 |
| Systematic Review and Meta-Analytics | 2 | 4.0 |
| Systematic Review | 2 | 4.0 |
| Prospective Cohort Study | 2 | 4.0 |
| Cross-Sectional Study | 1 | 2.0 |
| Case Report | 1 | 2.0 |
| **Total** | **50** | **100.0** |

When classified by evidence hierarchy according to the Oxford Centre for Evidence-Based Medicine (CEBM) framework (Table 6, Fig 7), Level 1 evidence (systematic reviews/meta-analyses and RCTs) comprised 12/50 (24.0%), Level 2 evidence (cohort studies) 7/50 (14.0%), Level 4 evidence (cross-sectional) 1/50 (2.0%), and Level 5 evidence (narrative/review articles and the case report) 30/50 (60.0%). Overall, the distribution is skewed toward lower-level evidence, reflecting the predominance of narrative and non-systematic reviews, with a smaller contribution from RCTs and formal evidence syntheses.

**Fig 7.**
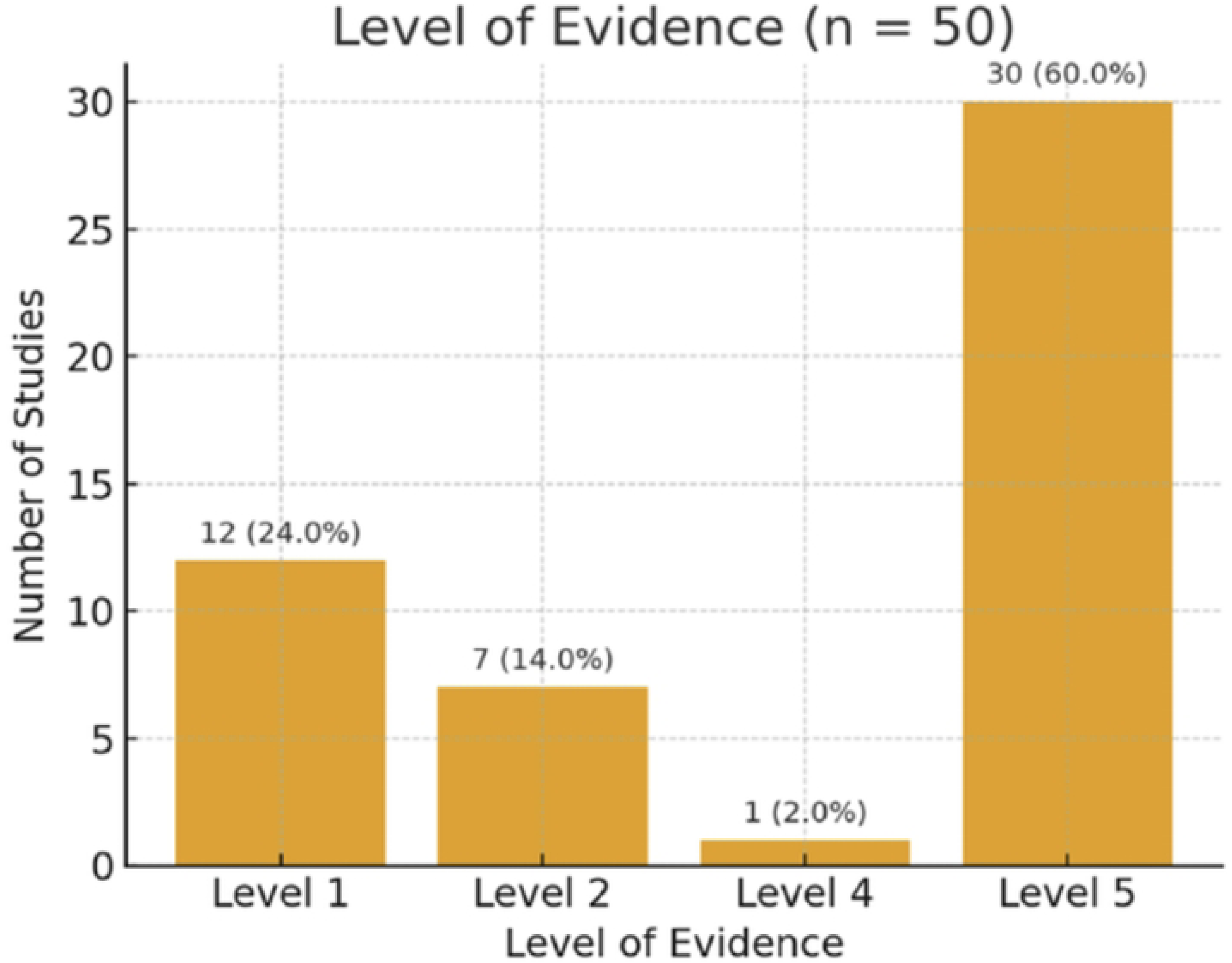
Levels of evidence represented in the top-cited CKD treatment studies.

**Table 6.**
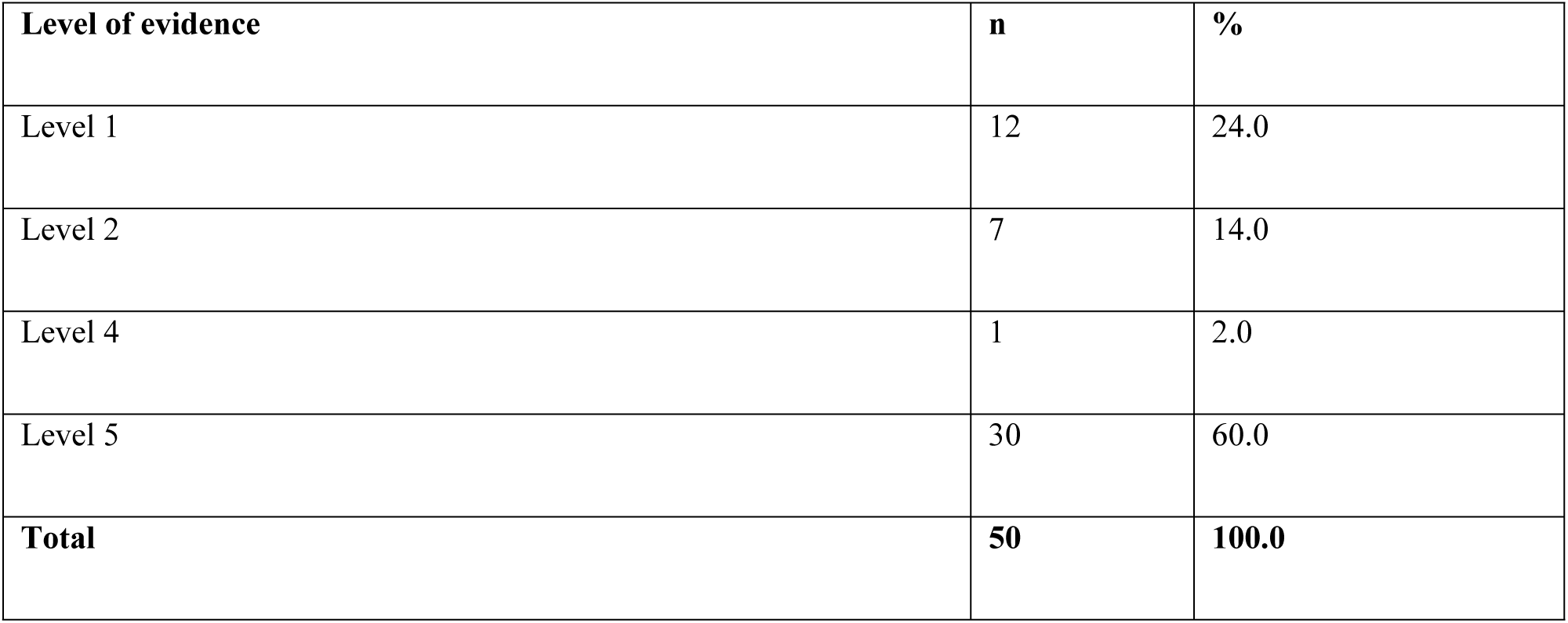
Level of Evidence Across the 50 Studies.

## Discussion

A bibliometric examination of the top 50 articles on the management of chronic kidney disease (CKD) reveals how a few seminal papers exert a substantial influence on clinical practice and research objectives. The field still mostly relies on retrospective observational research and narrative reviews, even though CKD is one of the major causes of morbidity and mortality globally^(5)^. Systematic reviews and meta-analyses received the most citations among the most cited papers, followed by randomized controlled trials; nevertheless, high-quality multicenter prospective studies were not as well-represented. This disparity suggests that while evidence synthesis has been useful in bringing information together, the groundwork for creative interventional research is somewhat limited.

The therapeutic themes found in the widely cited literature show how the field of therapy is changing. Strong scholarly influence was demonstrated by seminal papers that addressed renin-angiotensin system blockage, aldosterone antagonistic effects, and, more recently, sodium-glucose cotransporter 2 (SGLT2) inhibitors. An increasing trend toward both innovative pharmacological and integrative therapeutic approaches is also highlighted by randomized studies on bariatric surgery for obesity-related CKD and investigations of novel techniques such as gut microbiome manipulation and Chinese herbal medicine. The treatment of CKD has become increasingly complex, involving more extensive systemic and metabolic therapies in addition to traditional pharmaceutical approaches.

According to a geographic distribution analysis, low- and middle-income countries continue to provide relatively small contributions, whereas the majority of significant studies were produced in high-income regions like North America and Europe. The global application of these findings is called into question by this disparity, especially in regions with a high incidence of CKD but little resources. The ramifications are particularly pertinent in Saudi Arabia, where approximately 5% of people have CKD, most of them in stage 3. The primary causes are still diabetes, hypertension, and obesity, and therapy is made more difficult by medical issues such delayed diagnosis, dialysis dependence, and a lack of available transplants^(10,11)^. International findings cannot be fully incorporated into Saudi clinical and policy contexts due to the absence of localized bibliometric evaluations. Regarding methodological rigor, our results show that lower levels of evidence predominate. Only a small percentage of the top-cited literature included Level I and II evidence, such as systematic reviews, meta-analyses, and randomized controlled trials, but a significant fraction consisted of narrative reviews, expert opinions, and retrospective cohorts. Although clinical guidelines and policy have been greatly influenced by these evaluations, the lack of strong interventional research remains a major obstacle to strengthening the evidence base for treatments.

There are multiple limitations to this study. Historical precedence, self-citation, and journal impact all affect the number of citations. Furthermore, the search was restricted to English-language publications, which might have left out pertinent research that was written in other languages. In order to avoid language bias and guarantee wider coverage, it would be preferable for future analyses to incorporate studies in various languages.

In conclusion, there is still a dearth of original interventional evidence, and the therapeutic literature on CKD is mostly influenced by a few highly cited reviews and synthesis studies. Although bariatric surgery and SGLT2 inhibitors have become popular new treatments, they still require large multicenter randomized trials for additional validation. To ensure worldwide applicability, it is crucial to include more research from underrepresented locations. Integrating context-specific data with globally recognized research will be crucial for nations like Saudi Arabia to inform policy development and maximize treatment approaches.

## Conclusion

In conclusion, this bibliometric analysis highlights persistent imbalances in CKD treatment research, where academic influence remains concentrated in a limited number of narrative reviews and recurring therapeutic themes, with few multicenter interventional studies and minimal contributions from low-and middle-income countries (LMICs), including Saudi Arabia. Most of the top-cited publications emerged between 2010 and 2022 and were dominated by lower-level evidence primarily narrative and non-systematic reviews rather than high-quality randomized controlled trials or formal systematic syntheses.

Research clusters centered largely on RAAS blockade, SGLT2 inhibitors, and metabolic approaches, while alternative modalities such as traditional Chinese medicine, herbal interventions, and microbiome-targeted therapies occupied smaller, peripheral streams. The overall output remains heavily skewed toward high-income regions, raising concerns about the global applicability of current evidence, particularly in health systems with distinct epidemiologic profiles and resource limitations. Saudi Arabia exemplifies this imbalance: although CKD in the region is largely driven by diabetes, hypertension, and obesity, the nation’s limited research representation risks policy misalignment and underinformed guideline adaptation.

To our knowledge, this is the first bibliometric study focused on CKD treatment that integrates citation trends with geographic distribution and policy relevance, clearly identifying where the evidence is strong and where it remains critically lacking. Future research priorities should emphasize pragmatic multicenter RCTs, broader exploration of metabolic and nutritional interventions, and bibliometric assessments encompassing underrepresented regions and languages. Establishing coordinated trial networks within Saudi Arabia and the wider GCC, alongside strong enforcement of transparent reporting, will be essential to advance a more equitable, regionally relevant, and high-level evidence base in CKD care.

## Data Availability

All relevant data are within the manuscript and its Supporting Information files.

## Acknowledgment

None

## Supporting information

**S1 File. Dataset.** This file contains the full dataset used in the analyses.

